# Machine Learning Insights into HLA Noncoding Sequence Mismatches and Their Impact on DPB1 Matching in Hematopoietic Cell Transplantation

**DOI:** 10.1101/2024.09.13.24313580

**Authors:** Medhat Askar, Timothy L. Mosbruger, Grace Tzun-Wen Shaw, Haedong Kim, Yuncheng Duan, Andrew S. Allen, Jamie L. Duke, Timothy S. Olson, Dimitri S. Monos, Tristan J. Hayeck

**Affiliations:** QU Health and Department of Immunology, College of Medicine, Qatar University, Doha, Qatar; Division of Genomic Diagnostics, Department of Pathology and Laboratory Medicine, Children’s Hospital of Philadelphia, Philadelphia, Pennsylvania, USA; Department of Pathology and Laboratory Medicine, Perelman School of Medicine, University of Pennsylvania, Philadelphia, Pennsylvania, USA; Department of Biostatistics and Bioinformatics, Duke University, Durham, NC, USA

## Abstract

**Purpose:** HCT is vital for treating hematological malignancies, relying on HLA matching between unrelated patient-donor pairs to significantly reduce adverse outcomes. Recent studies recognize the potential impact of HLA-DPB1 mismatches on HCT outcomes. Multiple approaches focus on finding better-tolerated HLA-DPB1 mismatches. Additionally, recent studies suggest matching at noncoding HLA sequence may improve HCT outcomes. This study aims to evaluate different approaches for categorizing DPB1 mismatches in patient-donor pairs and explore the potential impact of noncoding mismatches (available in class I HLA genes) on clinical outcomes.

**Methods:** A retrospective study of 5,106 HCT pairs using Cox proportional hazards models, weighted by a machine learning algorithm, evaluates the impact of particular combinations of HLA-DPB1 mismatches in the context of noncoding HLA class I mismatches on outcomes of HCT. HLA-DPB1 mismatch criteria included T-cell epitope permissive/non-permissive mismatches, expression markers, and evolutionary clade mismatches.

**Results:** Two HLA-DPB1 mismatches, using multiple criteria, lead to significant hazards of acute graft versus host disease grades 2-4, in the T cell replete group. When HLA-DPB1 mismatches occurred across evolutionary clades (DP2 allele/low-expression patient vs DP5 allele/high-expression in the donor), the deplete group showed significant hazards for treatment-related mortality (TRM) (HR=1.94, p-value=8.9×10^-7^) and overall survival (OS) (HR=1.67, p-value=1.3×10^-5^) for additive effects of noncoding mismatches with two HLA-DPB1 mismatches.

**Conclusion:** Two HLA-DPB1 mismatches remain to predict worse outcomes. However, noncoding mismatches in HLA class I genes confer elevated hazards of TRM and OS in conjunction with mismatches across evolutionary branches of HLA-DPB1. Therefore, noncoding mismatches may inform donor selection in the presence of HLA-DPB1 mismatches and improve HCT outcomes, emphasizing the utility of comprehensive sequencing of HLA alleles in HCT settings.

## Introduction

Hematopoietic cell transplantation (HCT) stands as the sole curative option for numerous hematological malignancies, immunodeficiencies, and various disorders. Nonetheless, HCT carries a significant risk of acute graft-versus-host disease (aGVHD) and treatment-related mortality (TRM). Often, related donors are unavailable, necessitating the need for unrelated donors. The extent of HLA compatibility, between the donor and patient, is thought to induce divergent transplant-related immunologic responses.^1^ This, in turn, leads to clinical outcomes dependent on the relative level of HLA mismatches between the patient and donor. Therefore, the matching at HLA alleles in transplantation settings is pivotal for improving outcomes.

Matching patient-donor *HLA-DPB1* alleles is associated with improved HCT outcomes. However, HLA-DPB1 is separated from other HLA genes by a recombination hotspot ^2–5^, disrupting the linkage disequilibrium patterns in the region^6^ and complicating the ability to match *HLA-DPB1* between patients and donors when other HLA genes are matched (Figure 1E). Given the lower probability of *HLA-DPB1* matching, efforts have concentrated on approaches to find mismatches that are less deleterious to clinical outcomes.

**Figure 1.**
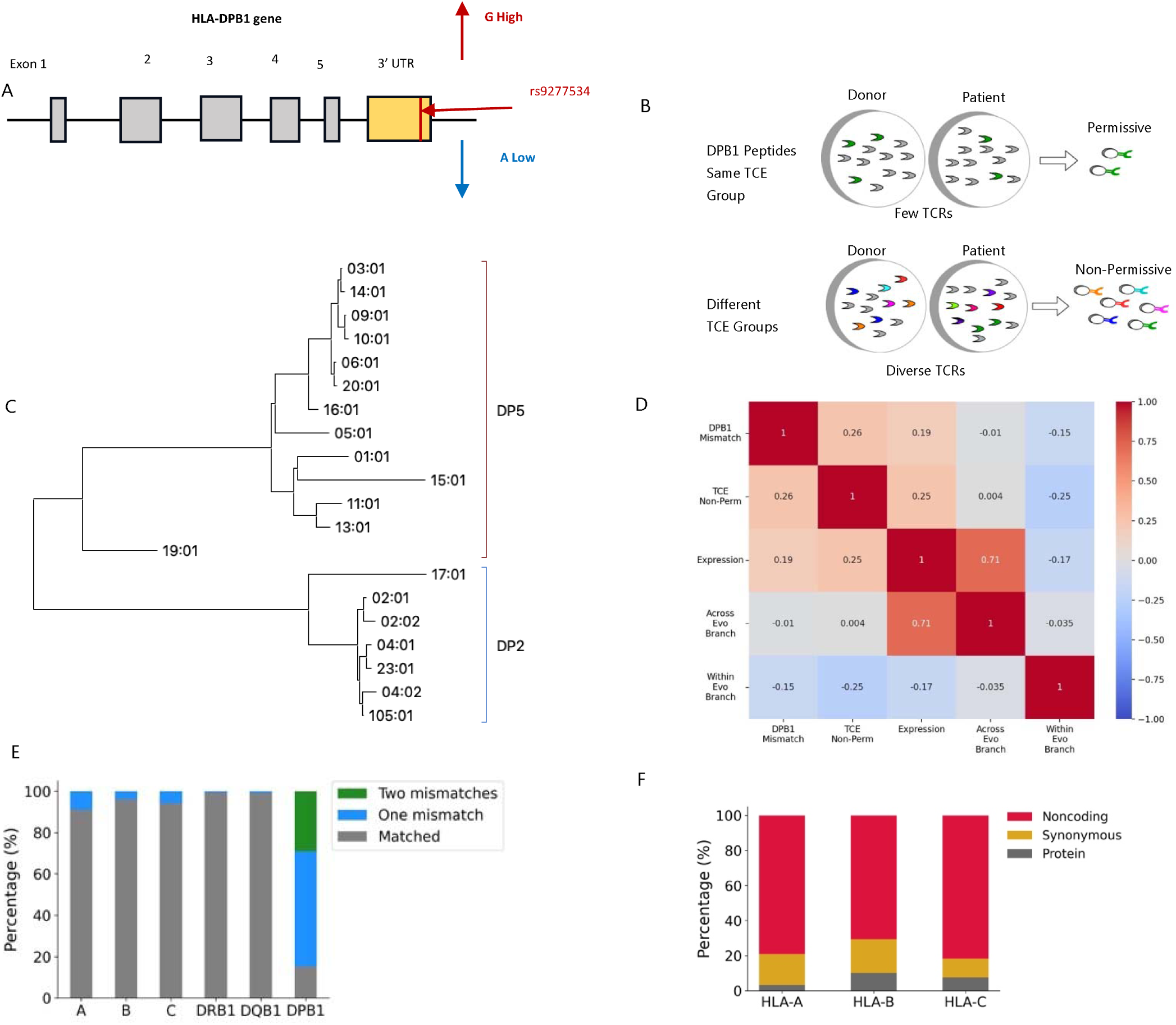
Mismatch models for DPB1 and frequency of mismatches by allele. Schematic representations of the different models for DPB1 mismatches comparing A) high versus low expression as tagged by expression marker rs9277534, B) TCE permissive versus non-permissiveness, and C) mismatches across evolutionary clades with a E) heat map comparing the relative correlation across the different models of DPB1 mismatches. Then looking at the D) relative percentages of either matched, one mismatch, or two mismatches at an allele level and then F) the percentages of either protein, synonymous or noncoding mismatches across class I HLAs.

Even before considering HLA-DPB1 matching, efforts to HLA match donors and recipients are primarily focused on matching across the coding regions (Figure 1F), especially within the antigen recognition domain (ARD). However, it has been reported that the extended haplotype context across the entire HLA genes, including noncoding mismatches, is also associated with HCT outcomes.^7,8^ For example, interactions between extended haplotypes involving HLA-DPB1 and MICA have been found to increase the risk of GVHD in HCT.^9^ Evidence suggests that regulatory elements residing within noncoding elements, such as microRNA located within HLA genes, likely have haplotype-specific effects on gene expression across the MHC region. ^10,11^ While it is believed that differential expression levels between patients and donors may influence HCT outcomes, as shown in studies of HLA-C, ^12^ ^10^ the broader impact of noncoding variations on HCT outcomes remains unclear.

Recently, in a cohort of unrelated HCT patient-donor pairs from the Center for International Blood and Marrow Transplantation (CIBMTR), Mayor et al.^13^ reported an improvement in clinical outcomes when HLA matching status was determined from ultra-high resolution (UHR) HLA genotyping [complete genomic characterization of class I genes (*HLA-A*, *-B* and *-C*); exons 2 and 3 for class II genes (*HLA-DRB1*, *-DQB1* and *-DPB1)*], showing the significance of moving beyond matching for ARD and including noncoding regions. Surprisingly and in contrast to earlier studies,^14^ there was a lack of association with worse outcomes when only HLA-DPB1 T-cell epitope (TCE) matching status was considered. However, a higher number of mismatches, at any locus, resulted in worse outcomes, where almost all mismatched pairs included at least one *HLA-DPB1* mismatch. Building off their findings, we therefore investigated the noncoding matching status in the context of different categories of DPB1 mismatches.

## Methods

### Study Design and Population

This is a retrospective study of a cohort of unrelated HCT patient donor pairs from the CIBMTR approved by the National Marrow Donor Program (NMDP) Institutional Review Board. The cohort studied has previously been described by Mayor et al. ^13^ The General Data Protection Regulation and internal CIBMTR regulations require that European Union subjects are excluded from datasets, therefore 32 European Union patients were excluded from this analysis. Additionally, two pairs were removed from the study because they were reported to have null alleles leaving 5,106 (Supplemental Table 1). The HLA typing utilized Pacific Biosciences’ single-molecule real-time DNA sequencing method, for class I genes (HLA-A, B, and C) this included all exons, introns and untranslated regions, and class II genes (HLA-DRB1, DQB1 and DPB1) included exons 2 and 3.

#### Defining different Criteria for DPB1 Mismatches

This study examines multiple methods of classifying DPB1 mismatches in the context of HCT including: any DPB1 mismatches, TCE permissive versus nonpermissive, high versus low expression mismatches, and evolutionary branch mismatches across DP2 and DP5 clades. The simplest scheme is to count any DPB1 mismatches between patient-donor pairs without imposing any additional criteria.

For matching based on TCE permissiveness (Figure 1B), the algorithm of Crivello et al ^15^ was utilized. For TCE permissiveness, DPB1 mismatches between patient and donor may induce alloreactive T-cell responses, based on the cross reactivity of different DPB1 defining the classification as permissive or non-permissive TCE mismatches.^15–20^ Another approach is mismatches of DPB1 expression levels, where expression was defined using the imputed rs9277534 SNP which has been reported to be strongly associated with DPB1 expression levels being high or low.^21^ The expression allele was imputed by matching the reported DPB1 typing with previously reported exon 2, exon 3 and rs9277534 linkages.^22^ DPB1 alleles containing exon 2 and 3 sequences without known rs9277534 linkages were set to unknown (n=17). Expression mismatch coding was defined such that if the patient has the imputed rs9277534-G allele and has any mismatch across DPB1 with the donor then they are considered high expression mismatches. If the patient has two rs9277534-A alleles and there is any mismatch across DPB1 between the patient and donor, then the patient is considered to be a low expression mismatch.^23^ This is independent of the donor’s expression markers.

The study also investigated an evolutionary model using mismatches across evolutionary branches (Figure 1C),^24,25^ specifically DP2 and DP5. This is intricately linked to the expression model, as individuals with alleles in the HLA-DP2 clade consistently contain the rs9277534-A allele (low expression marker), while those in the HLA-DP5 group predominantly have the rs9277534-G allele (high expression marker). These models, expression and evolutionary clades, exhibit a degree of overlap in their matches or mismatches (Figure 1D), primarily due to robust positive linkage disequilibrium, particularly in the exon 3 to 3’-untranslated region, encompassing rs9277534. For simplicity, in this paper, we will commonly refer to this criterion as the DP2 and DP5 clades and the evolutionary model. However, it’s crucial to note that this is essentially equivalent to searching for matches across patient-donor expression allele markers, rs9277534, and as such, direct inferences regarding mechanistic implications cannot be made. The focus is instead on the overall predictive utility for clinical guidelines in different matching strategies.

In settings where the analysis considered two DPB1 mismatches, the category indicates at least one mismatch in the group. For example, when restricting to two DPB1 mismatches when looking at patient having at least one allele in the DP2 clade versus the donor having at least one allele in the DP5 clade would be coded as DP2vsDP5. This is similar for expression high/low coding. For TCE permissiveness, we look across the combined set of DPB1 alleles for the patient and donor, and if any non-permissive mismatches occur, they are categorized as nonpermissive, otherwise if they only have permissive mismatches, the pair evaluates as permissive.

#### Defining noncoding mismatches

Noncoding mismatches are defined as alleles that differ in their intronic or UTR sequences for HLA class I alleles or any synonymous substitution in exons of any of the six genes being tested. The mismatch count is based upon the total number of alleles with mismatches (i.e., any difference in the 3^rd^ or 4^th^ field), as opposed to the number of nucleotide differences.

## Statistical Methods

For analysis, the population was first split into two cohorts of individuals that received in vivo T-cell-depleted transplants (n = 2,030) who received anti-thymocyte globulin (ATG) or Alemtuzumab and T-Cell-repleted transplants (n = 3,076). Interventions like ATG are used to prevent GVHD in settings of HCT. However, T cell depletion can also compromise alloimmune and graft versus leukemia response and in turn affect survival^26^ which may be relative to patient– donor mismatch status. Therefore, repletion or depletion acts as a moderator of the effect of relative HLA mismatches, which means it’s expected that receiving a therapeutic, ATG or Alemtuzumab, may impact the severity of outcomes and relative impact of level of mismatches.^13^ For these reasons, we stratified the analysis into the replete and deplete cohorts.

A series of weighted cox proportional hazards models were fit using weighting based on propensity scores (PS) estimates to control for potential confounders.^27–30^ Propensity scores in this context represent the probability of finding a UHR match, determined by the patient’s characteristics. They help balance the influence of the various characteristics (Supplemental Table 1) between patient-donor pairs who receive an exact match and those who do not.^31,32^ To calculate the propensity scores a gradient boosted model (GBM), which is a machine learning algorithm, was fit (Supplemental Figure 1) using the R (version 4.3.1) twang (version 2.5) software.^31,32^

The GBM algorithm operates by iteratively building a sequence of predictive models, or decision trees, and then combining them into an ensemble model. This process aims to minimize prediction errors while controlling overfitting and variability. Each new model is trained to correct the errors made by the previous models, leading to gradual improvement in predictive performance. To manage overfitting and enhance stability, a learning rate (or shrinkage factor) is used. This factor scales the contribution of each decision tree, helping to reduce variability and refine the ensemble model. GBM offers advantages over other approaches such as stepwise regression or penalized regression, which involve variable selection. These methods risk biasing exposure effect estimates by potentially eliminating covariates that influence exposure selection.^41^ In contrast, other methods like stepwise regression or penalized regression are less flexible and may inaccurately define the relationship between covariates and exposure selection when compared to decision tree-based models such as GBM. Instead of selecting a single model with a particular set of covariates GBM allows for an aggregate across a range of models. Here the GBM assumes there are no unmeasured confounders and no time varying confounding when applied in such settings, as would these other similar approaches. The objective of fitting this type of model is not to interpret the effects of these covariates but to optimally control for them, thereby improving the inference on the main exposure of interest: HLA mismatches.

The GBM was fit with 5,000 trees, a shrinkage parameter of 0.01, an interaction depth of 3 using the absolute standardized mean difference as the stopping criteria for the balance. For the estimand, Average Treatment effect in Treated (ATT) was chosen as opposed to the Average Treatment Effect (ATE). The ATE is typically of more interest if every treatment or exposure maybe potentially offered to every member the population^39,40^ whereas in transplantation settings if a perfect match (the exposure) is both known and available that would be chosen–so ATT was chosen instead.

After controlling for covariates, the main objective is to assess HLA mismatches. In situations where testing was conducted to assess the impact of one or more alleles with noncoding mismatches (in the context of DPB1 mismatches) compared to baseline categories, the model assumed additive effects so it will be referred to as the additive model. In simpler terms, this means that each additional noncoding mismatched allele is considered to incrementally increase the hazards of adverse outcomes by the model. Using a conservative p-value threshold of 1% with adjustment for multiple hypothesis, eight criteria of matching, gives a p-value threshold of 1.25 × 10^-3^.

In the section *The Influence of Accounting for Noncoding Sequences*, subsequent analysis was performed to assess the influence of noncoding mismatches (3^rd^ or 4^th^ field differences) in the context of DPB1 mismatches. For this purpose, tests were performed to compare hazard models with and without including noncoding mismatch information. Analysis was performed exclusively focused on protein coding, mirroring two-field typing by concealing established noncoding sequences then that was contrasted with analysis integrating noncoding sequencing (four-field typing). The section does not aim to determine significant associations. The aim is to assess whether the absence of noncoding variation introduces bias (shifts the hazard ratios) or inflates results (changes the p-values and relative significance) by omitting noncoding mismatches. For illustrative purposes, a p-value threshold of 1% will be applied.

## Results

### Comparison of HCT outcomes when using different DPB1 matching criteria

Analysis was performed comparing the different matching strategies (DPB1 allele mismatch, DPB1 TCE permissive mismatches, DPB1 expression status, and DPB1 evolutionary clades) across all studied outcomes (aGVHD, OS, TRM) when the pair is mismatched for DPB1 (restricting to 10/10). Although a general trend of increased hazard of aGVHD was observed when patient-donor pairs have only one DPB1 mismatch as compared to the baseline model of completely matched cases (12/12 UHR matched), none of the DPB1 matching strategies resulted in outcomes with significant differences (Supplemental Table 2-Supplemental Table 4). However, when we considered pairs with two DPB1 mismatched alleles and focused on the replete cohort, significant hazards of aGVHD grades 2 – 4 (α < 1.25 × 10^-3^) were observed in the following conditions: any two DPB1 mismatches (HR = 1.40, p-value = 7×10^-4^), TCE non-permissive mismatches (HR = 1.46, p-value = 5×10^-4^), patient DP5 versus donor DP2 mismatches (HR = 1.54, p-value = 7×10^-4^), and high expression (HR = 1.43, p-value = 1.1×10^-3^) were all significant for the replete cohort (Table 1 and Figure 2). Notably, when considering a more **severe aGVHD status (grades 3 – 4), only TCE non-permissive mismatches maintained significant hazards (HR = 1.76, p-value = 9×10^-4^)** (Supplemental Table 5).

**Figure 2.**
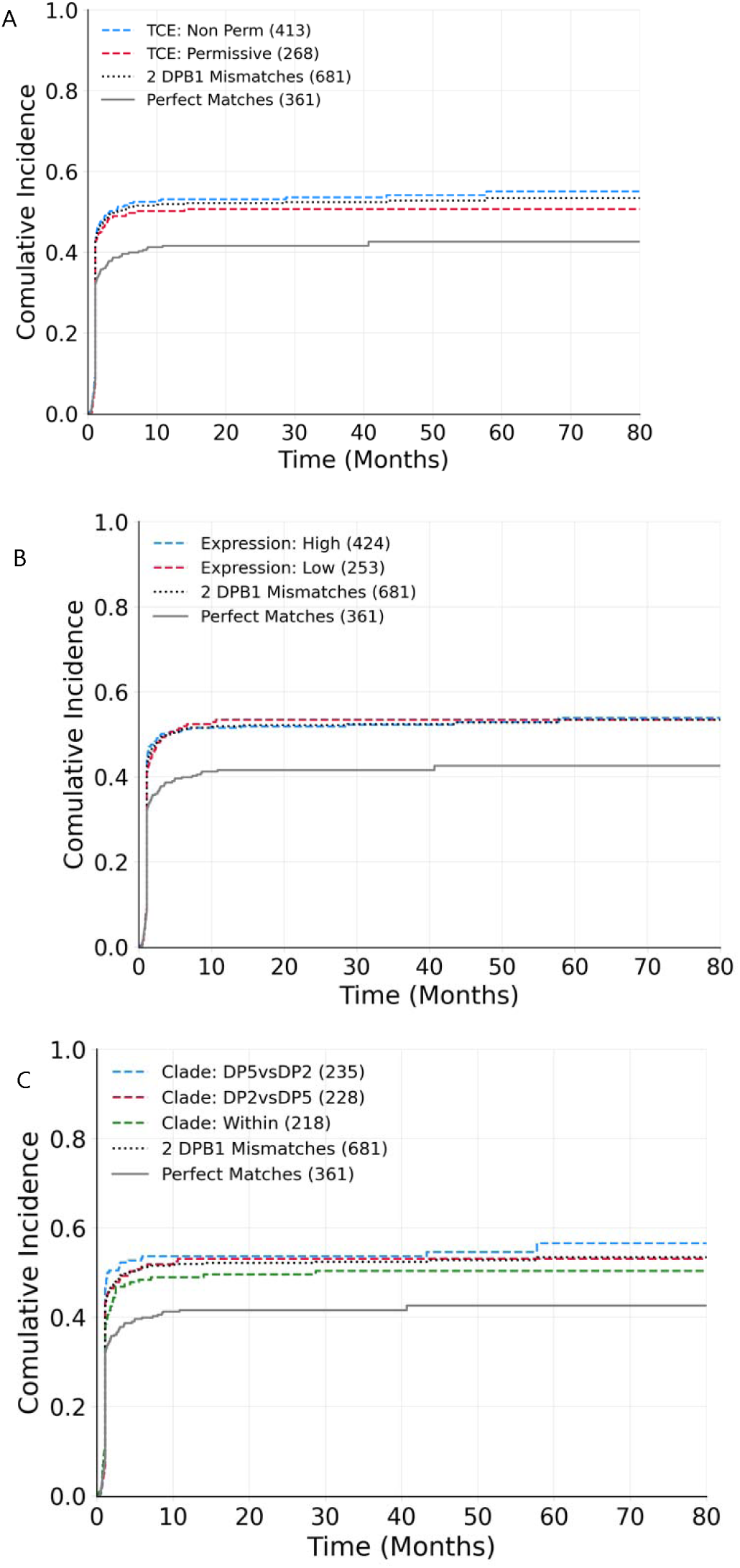
Hazards of aGVHD grades 2-4 in the context of two DPB1 mismatches comparing scenarios of DPB1 mismatches for replete subgroup. For all plots, the cumulative incidence of aGVHD is shown for the baseline group with perfect HLA matching (gray solid line) and with any 2 DPB1 mismatches (gray dotted line). These are then compared to pairs having two DPB1 mismatches with either A) T-cell epitope (TCE) permissive (red) versus non-permissive mismatches (blue), B) high (blue) versus low (red) expression based on the rs9277534 expression marker, and C) mismatches across evolutionary clades, DP5vsDP2 (blue) or DP2vsDP5 (red), and mismatches within the same evolutionary clade (green).

**Table 1.**
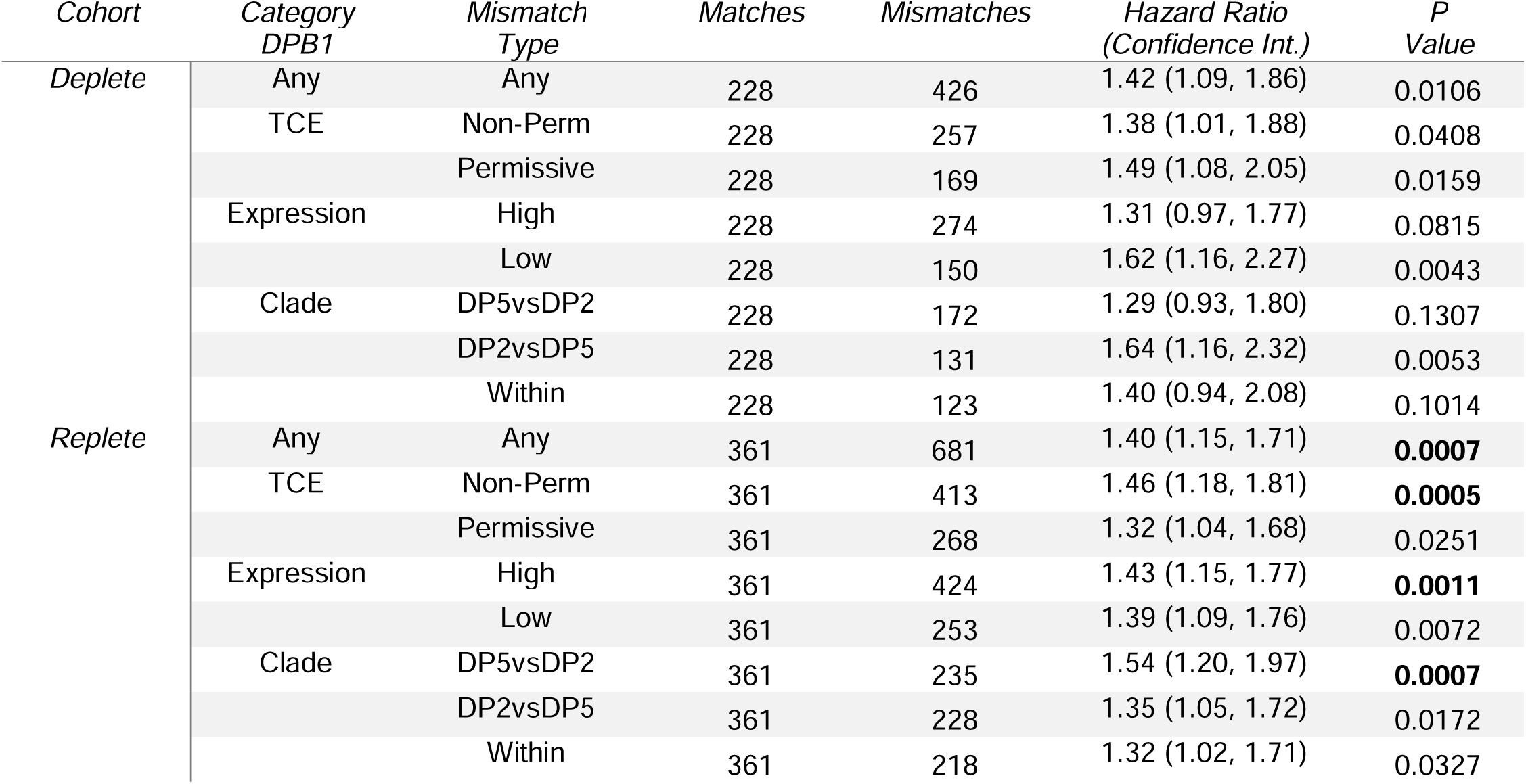
Hazards of aGVHD grades 2-4 for pairs having 2 DPB1 mismatches (restricting to 10/10 matching on other alleles) versus complete matching (12/12). The results are split first by the deplete and replete cohorts, then a comparison across DPB1 mismatch categories: 1) any type of DPB1 allele mismatches (any), 2) TCE permissiveness, 3) patient DPB1 expression, and 4) evolutionary clade. For the evolutionary model DP5vsDP2 corresponds to a patient with DP5 versus a donor with DP2 and is flipped for DP2vsDP5. For expression profiles, high expression signifies the presence of the high expression marker, rs9277534-G, in the patient (GG or AG), while low expression indicates the presence of only the low expression marker, rs9277534-A, in the patient (AA). In both scenarios, the expression category is assigned regardless of the expression level of the donor.

| Cohort | Category<br>DPB1 | Mismatch<br>Type | Matches | Mismatches | Hazard Ratio<br>(Confidence Int.) | P<br>Value |
| --- | --- | --- | --- | --- | --- | --- |
| Deplete | Any | Any | 228 | 426 | 1.42 (1.09, 1.86) | 0.0106 |
|  | TCE | Non-Perm | 228 | 257 | 1.38 (1.01, 1.88) | 0.0408 |
|  |  | Permissive | 228 | 169 | 1.49 (1.08, 2.05) | 0.0159 |
|  | Expression | High | 228 | 274 | 1.31 (0.97, 1.77) | 0.0815 |
|  |  | Low | 228 | 150 | 1.62 (1.16, 2.27) | 0.0043 |
|  | Clade | DP5vsDP2 | 228 | 172 | 1.29 (0.93, 1.80) | 0.1307 |
|  |  | DP2vsDP5 | 228 | 131 | 1.64 (1.16, 2.32) | 0.0053 |
|  |  | Within | 228 | 123 | 1.40 (0.94, 2.08) | 0.1014 |
| Replete | Any | Any | 361 | 681 | 1.40 (1.15, 1.71) | **0.0007** |
|  | TCE | Non-Perm | 361 | 413 | 1.46 (1.18, 1.81) | **0.0005** |
|  |  | Permissive | 361 | 268 | 1.32 (1.04, 1.68) | 0.0251 |
|  | Expression | High | 361 | 424 | 1.43 (1.15, 1.77) | **0.0011** |
|  |  | Low | 361 | 253 | 1.39 (1.09, 1.76) | 0.0072 |
|  | Clade | DP5vsDP2 | 361 | 235 | 1.54 (1.20, 1.97) | **0.0007** |
|  |  | DP2vsDP5 | 361 | 228 | 1.35 (1.05, 1.72) | 0.0172 |
|  |  | Within | 361 | 218 | 1.32 (1.02, 1.71) | 0.0327 |

When the deplete cohort was studied with two DPB1 mismatched alleles, no DPB1 matching criteria showed significant hazards for aGVHD grades 2 – 4 (Table 1). However, patient DP2 versus donor DP5 and DPB1 low expression mismatches were marginally above the alpha cutoff threshold of 1.25 × 10^-3^ (Table 1). No other hazards were significant across the other testing outcomes when considering the different DPB1 matching strategies (Supplemental Table 6 and Supplemental Table 7).

### Noncoding Mismatches in the context of DPB1 Mismatches

The following analysis accounts for the role of noncoding mismatches in HLA class I genes in the context of the different categories of DP matching on different outcomes. First, analysis was done to isolate if noncoding mismatches (comparing against a baseline of 12/12 matched pairs) alone were associated with HTC outcomes. On their own, noncoding mismatches did not have any significant effect on outcomes.

Next, we investigated the scenario of having one DPB1 mismatched allele and additive effects of noncoding mismatches in the other HLA genes, compared to the baseline of pairs with a single DPB1 allele mismatch that is also 10/10 UHR matched. For all cases, no significant hazard ratios for any of the primary outcomes of interest (Supplemental Tables 8 – 10).

Next, we consider the effect of noncoding mismatches in the context of cases that possess two DPB1 allele mismatches. Here, the baseline for these analyses are cases that have two DPB1 allele mismatches and are also 10/10 UHR matched. Two different types of models were constructed, first a model comparing cases that have a single allele with a noncoding mismatch outside of HLA-DPB1, compared to baseline, and second an additive model that compares the incremental effects of having additional HLA alleles with noncoding mismatches in the non-DPB1 genes compared to the baseline. Neither of these models identified significant hazards for TRM (Table 2, single allele with a noncoding mismatch model; Table 3, additive model), OS or aGVHD (data not shown) for the replete cohort.

**Table 2.** Hazard ratios of treatment related mortality between pairs having one noncoding mismatch (9/10) and two DPB1 mismatches versus a baseline of pairs that are 10/10 matched with two DPB1 mismatches. The purpose is to restrict testing for the hazards of TRM within DPB1 mismatch category plus one noncoding mismatch relative a baseline of no noncoding mismatches (10/10 matching) with two DPB1 mismatches.

| Cohort | Category<br>DPB1 | Mismatch<br>Type | Matches | Mismatches | Hazard Ratio<br>(Confidence Int.) | P-Value |
| --- | --- | --- | --- | --- | --- | --- |
| Deplete | Any | Any | 446 | 102 | 1.38 (0.91, 2.10) | 0.1265 |
|  | TCE | Non-Perm | 446 | 68 | 1.46 (0.94, 2.26) | 0.0919 |
|  |  | Permissive | 446 | 34 | 1.22 (0.54, 2.73) | 0.6317 |
|  | Expression | High | 446 | 61 | 1.37 (0.83, 2.27) | 0.2227 |
|  |  | Low | 446 | 40 | 1.40 (0.75, 2.61) | 0.2894 |
|  | Clade | DP5vsDP2 | 446 | 37 | 1.17 (0.60, 2.27) | 0.6472 |
|  |  | DP2vsDP5 | 446 | 38 | 2.42 (1.47, 3.99) | **0.0005** |
|  |  | Within | 446 | 27 | 0.64 (0.24, 1.73) | 0.3819 |
| Replete | Any | Any | 715 | 136 | 0.69 (0.46, 1.04) | 0.0781 |
|  | TCE | Non-Perm | 715 | 81 | 0.74 (0.44, 1.23) | 0.2472 |
|  |  | Permissive | 715 | 54 | 0.64 (0.35, 1.18) | 0.1549 |
|  | Expression | High | 715 | 80 | 0.70 (0.42, 1.19) | 0.1859 |
|  |  | Low | 715 | 55 | 0.69 (0.38, 1.25) | 0.2230 |
|  | Clade | DP5vsDP2 | 715 | 40 | 0.57 (0.26, 1.26) | 0.1644 |
|  |  | DP2vsDP5 | 715 | 56 | 0.79 (0.45, 1.40) | 0.4224 |
|  |  | Within | 715 | 40 | 0.66 (0.32, 1.37) | 0.2686 |

**Table 3.**
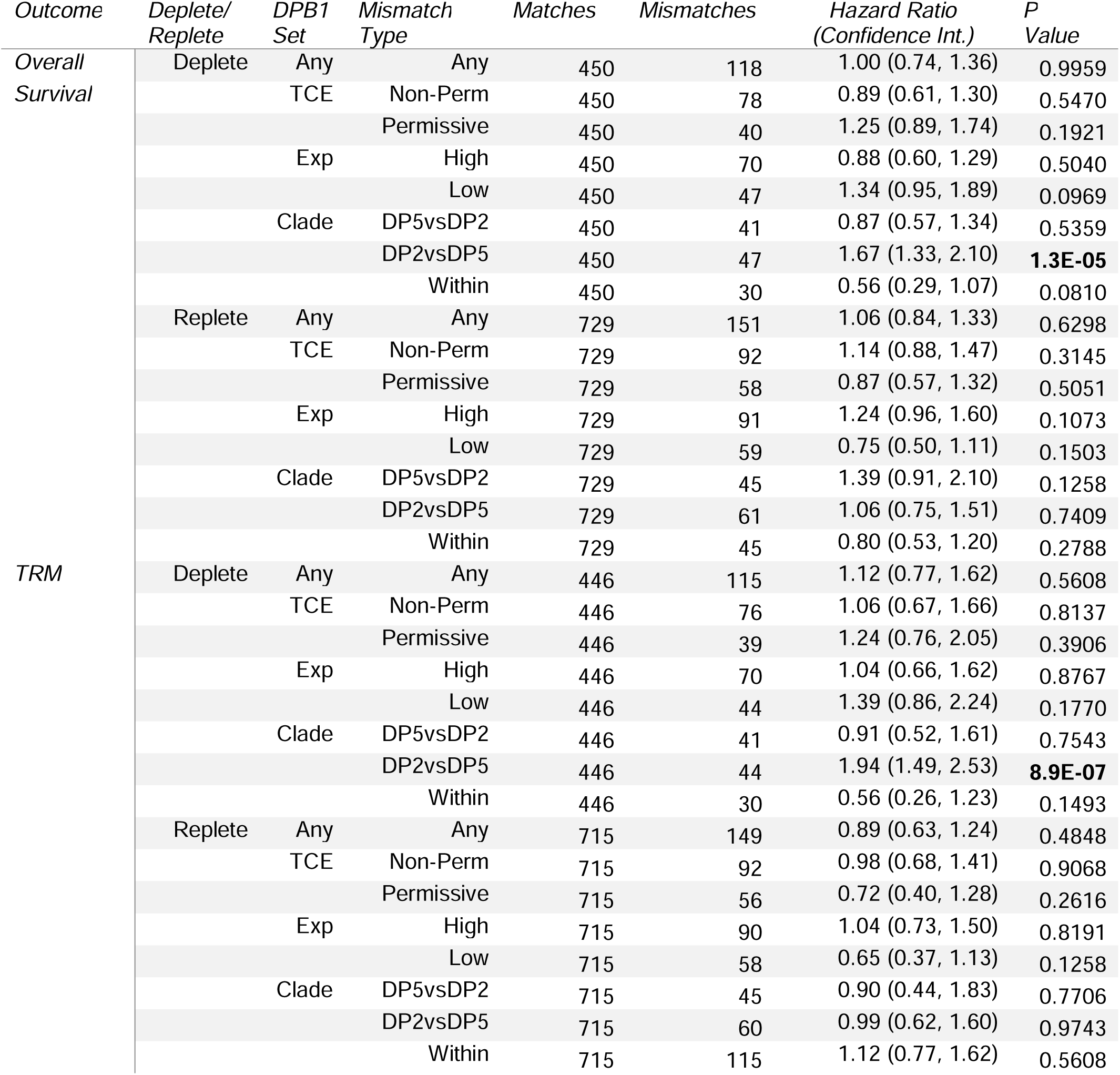
Overall survival and treatment related mortality hazard ratios for the additive model of noncoding mismatches with 2 DPB1 mismatches versus baseline of no noncoding mismatches (10/10 matching) with 2 DPB1 mismatches. For each category of DPB1 mismatches there is at least one DPB1 mismatch in the corresponding category.

However, significant hazards are observed in these two types of models when the deplete cohort is analyzed. In this cohort, we found a significant hazard of TRM when the patient is 9/10 UHR matched and two DPB1 allele mismatches, where the patient’s DP2 clade allele is mismatched against the donor’s DP5 clade allele (HR=2.42, p-value=5.1×10^-4^; Table 2). Furthermore, when additional noncoding mismatched alleles outside of the HLA-DPB1 gene are included in the analysis as part of the additive model, significant hazards are found in this same DP mismatch scenario (patient DP2 vs donor DP5) for both TRM (HR=1.94, p-value = 8.9×10^-7^) and OS (HR=1.67; p-value = 1.3×10^-5^) (Table 3 and Figure 3A and 3B).

**Figure 3.**
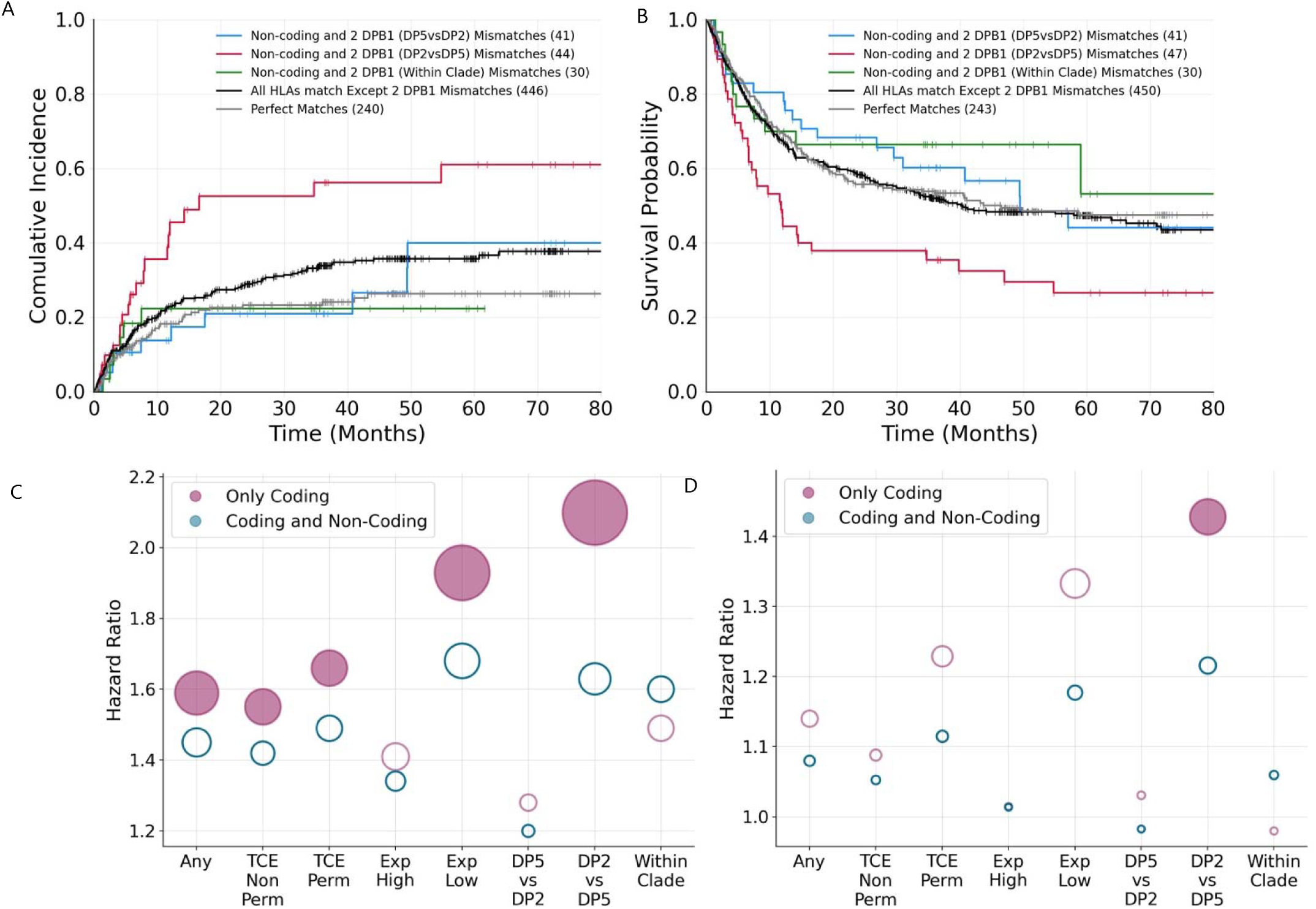
Hazards of treatment related mortality and overall survival in the context of two DPB1 evolutionary clade mismatches and noncoding mismatches in HLA class I alleles for the deplete cohort. The top panels compare A) TRM cumulative incidence and B) overall survival rates for different mismatch categories: 12/12 matches (gray), 2 DPB1 mismatches with 10/10 matching elsewhere (gray), and three categories where pairs have two DPB1 clade mismatches with one or more noncoding mismatches with 10/10 matching. The lower plots show the hazard ratios for C) TRM and D) OS, comparing 12/12 matches versus 10/10 matches with 2 DPB1 MM. Pink corresponds to matches accounting for only coding sequences (two field typing, not considering the status of noncoding mismatches); blue corresponds to matches of both coding and noncoding regions (whereby donor and recipient do not have noncoding mismatches). The hazard ratio dot sizes are proportional to the -log_10_(p-values), larger dots have smaller p-values and filled in dots are significant at a threshold of 0.01 or lower. Higher hazards indicate worse outcomes relative to baseline.

Since these results were highly significant, additional testing was performed in the subset of cases having patient DP2 vs donor DP5 mismatch, and comparing a baseline of two DPB1 allele mismatches versus an additive model incorporating noncoding mismatches. **When comparing to a baseline within category patient DP2 vs donor DP5, significant hazards remain for the additive effect of noncoding variants for both TRM (HR=1.82, p-value = 1×10^-4^) and OS (HR=1.54; p-value = 1.2×10^-3^).**

### The Influence of Accounting for Noncoding Mismatches

Noncoding mismatches were typically unavailable in previous studies linking DPB1 allele mismatches with HCT outcomes, potentially conflating their effects with DPB1 mismatches. To disentangle the two elements at play, we analyzed different DPB1 mismatch categories and assessed whether noncoding mismatches bias or inflate the significance of DPB1 mismatches in HCT outcomes. We used two baseline models for 12/12 matches: one considering only coding-level matches (2-field typing) and another including both coding and noncoding regions (4-field typing). Consistent bias was observed in the deplete cohort, whereby the hazard ratios were inflated when only coding mismatches were considered for both TRM (upward bias up to 0.47 in hazards; Supplemental Table 11) and OS (upward bias up to 0.21 in hazards; Supplemental Table 12), except for within clade mismatches. The bias pattern was reversed for the replete cohort, showing a downward bias in TRM (downward bias up to 0.17 in hazards; Supplemental Table 11) and OS (downward bias up to 0.11 in hazards; Supplemental Table 12), except for DP2 vs DP5 (HR biased upward of 0.07) and high expression (negligible upward bias of 4×10^-3^) (Supplemental Figure 2).

Next, we investigated how noncoding allele mismatches would impact the interpretation of the statistical significance of different DPB1 mismatch models with outcomes (Supplemental Tables 15 and 16). No DP mismatch categories were significant for the replete cohort and were not considered further. For the deplete group, many of the DP mismatch categories have significant hazards for TRM when compared to a baseline that is 12/12 matched for coding sequences only (Figure 3C and D, pink filled dot for the categories of any, TCE non-permissive, TCE permissive, Low expression, and DP2 vs DP5). However, when the baseline is changed to 12/12 matched for coding and noncoding sequences no category is significant for TRM (Figure 3C). A similar trend is observed for OS, whereby DP2 vs DP5 is the only significant category with the baseline of 12/12 matched for coding sequences but loses significance when the baseline is 12/12 matched for coding and noncoding sequences.

## Discussion

Novel technologies are making comprehensive sequencing more cost-effective, raising the question of whether detailed sequencing of HLA loci and the broader MHC could improve HCT outcomes. This study first isolated different models of DPB1 mismatches comparing 10/10 matching with complete matching at HLA alleles (12/12), demonstrated that having two DPB1 mismatches results in worse hazards of aGVHD, while the relative influence of the specific mismatch model (ie any, non-permissive, expression, or clade) may be irrelevant. Worse outcomes with multiple DPB1 mismatches aligns with both Wang^33^ and Petersdorf’s^23^ findings. This does not directly contradict Mayor’s work, ^13^ whereby Mayor reported that TCE nonpermissive mismatches were not significantly associated with worse outcomes. Mayor et al. treated DPB1 mismatches as either TCE permissive or non-permissive, independent of the number of DPB1 mismatched alleles (n = 1 or 2), which dilutes the effect we observe when there are two DPB1 mismatched alleles. Beyond the association between two HLA-DPB1 mismatches and aGVHD there were notable differences across the deplete and replete cohorts. In the deplete group, although none of the hazards reached significance at the adjusted threshold, the impact of DPB1 mismatches appears reversed compared to the replete cohort (Table 1) with patient DP2 versus donor DP5 and DPB1 low expression mismatches marginally above significance threshold. Specifically, higher hazards for aGVHD 2-4 were observed in cases of permissive, low expression, and patient DP2 versus DP5 donor, directly contrasting the replete counterpart.

Next, noncoding mismatches were considered. Noncoding mismatches alone did not have any significant effect on outcomes. Since so few patient-donor pairs have exclusively noncoding mismatches, this study would be severely under powered to detect any direct effect of noncoding mismatches (if they exist), similar to what is noted by Hurley.^34^ Previous findings have shown worse outcomes with nonpermissive mismatches in the context of another mismatch,^35^ these studies were restricted to mismatches across coding regions, where UHR testing was likely not available. This study was on the same cohort and genotyping used in the work of Mayor et al. ^13^ Our findings are complimentary to that of Mayor’s, ^13^ where they found higher number mismatches tended to result in worse HCT outcomes, irrespective of being coding or noncoding mismatches. However, their study had a different focus, they did not explicitly test for the impact of noncoding mismatches specifically nor did they consider different types of DBP1 mismatches. Our findings show that, in specific settings involving certain DPB1 mismatches, an increase in noncoding mismatches is associated with significant hazards for both TRM and OS. In the subgroup of DP2 versus DP5 with two DPB1 mismatches, the addition of a single noncoding mismatch had significantly worse outcomes, within the deplete cohort (Table 2). The significant hazards of TRM and OS in the additive noncoding model (Table 2-Table 3 and Figure 3) leaves uncertainty regarding whether the underlying disruptive mechanism corresponds to evolutionary factors, expression, or protein structure of linked genes. All possibilities are plausible. Further studies with larger sample sizes, whereby all classical HLA genes are fully characterized (including all intronic, exonic and untranslated regions), and functional follow-up of clinical outcomes are required to elucidate the true underlying causes.

The follow-up analysis comparing only the coding mismatches verses including noncoding mismatches showed both biases (generally hazards appear worse) and higher significance for both TRM and OS in the deplete group when noncoding mismatches are missing (Figure 3C and D). This potentially explains why some studies previously found that DPB1 mismatches by themselves account for significant hazards of TRM or worse OS. By attributing the effect solely to DPB1 mismatches, it is unclear whether the signal is derived from the DPB1 mismatches themselves or the noncoding mismatches elsewhere.

Our findings support prior studies that report HLA haplotype mismatches/heterogeneity negatively impacting HCT outcomes and augment the impact of mismatches at HLA-DPB1 locus. We hypothesize that matching for noncoding sequence is a marker of haplotype homogeneity and consequently noncoding mismatches may augment the impact of mismatches at other HLA loci ^36^ or more broadly across the MHC.^7,9,37,38^ Furthermore, it is also possible that the noncoding mismatches have regulatory impacts or are in linkage with other variants within the MHC haplotype, that could in turn effect immune related genes and HCT outcomes.

In conclusion, this study confirmed that having multiple HLA-DPB1 mismatches is associated with aGVHD and independently found that elevated hazards of both TRM and OS were associated with mismatches across evolutionary branches (or expression differences) of HLA-DPB1 in conjunction with an additive effect of noncoding mismatches in class I HLA genes. Analyzing the impact of the availability of noncoding mismatches shed light on why previous studies may have worse hazards of TRM or OS solely attributed to DPB1 mismatches. Incorporating noncoding sequences has the potential to enhance our understanding of additional attributes to the deleterious effect of mismatches at different HLA loci, inform HCT donor selection and ultimately improving clinical outcomes in HCT.

## Data Availability

All data produced in the present work are contained in the manuscript.

## Supplement

**Supplemental Table 1.** Summary of characteristics of patients and donors.

| Variable | Overall | T-Cell-Depleted<br>(ATG or Campath) | T-Cell-Replete |
| --- | --- | --- | --- |
| Number of patients | 5 106 | 2030 | 3076 |
| Primary disease for HCT |  |  |  |
| AML | 2551 (50) | 1042 (51) | 1509 (49) |
| ALL | 800 (16) | 285 (14) | 515 (17) |
| MDS | 1755 (34) | 703 (35) | 1052 (34) |
| Disease status at transplant |  |  |  |
| Early | 2384 (47) | 959 (47) | 1425 (46) |
| Intermediate | 690 (14) | 263 (13) | 427 (14) |
| Advanced | 1963 (38) | 776 (38) | 1187 (39) |
| Missing | 69 (1) | 32 (2) | 37 (1) |
| Recipient age group |  |  |  |
| 0-10 | 152 (3) | 81 (4) | 71 (2) |
| 11-18 | 135 (3) | 64 (3) | 71 (2) |
| 19-29 | 400 (8) | 165 (8) | 235 (8) |
| 30-39 | 432 (8) | 158 (8) | 274 (9) |
| 40-49 | 702 (14) | 256 (13) | 446 (14) |
| 50-59 | 1197 (23) | 472 (23) | 725 (24) |
| 60+ | 2088 (41) | 834 (41) | 1254 (41) |
| Median (range) | 57 (1-84) | 57 (1-84) | 57 (1-81) |
| Recipient sex |  |  |  |
| Male | 2909 (57) | 1185 (58) | 1724 (56) |
| Female | 2197 (43) | 845 (42) | 1352 (44) |
| Race |  |  |  |
| White | 4560 (89) | 1816 (89) | 2744 (89) |
| Other | 458 (9) | 181 (9) | 277 (9) |
| Missing | 88 (2) | 33 (2) | 55 (2) |
| Donor age group, years |  |  |  |
| 19-29 | 3 175 (62) | 1289 (63) | 1886 (61) |
| 30-39 | 1153 (23) | 449 (22) | 704 (23) |
| 40-49 | 596 (12) | 235 (12) | 361 (12) |
| 50-59 | 178 (3) | 57 (3) | 121 (4) |
| 60+ | 4 (<1) | 0 (<1) | 4 (<1) |
| Median (Range) | 28 (18-61) | 28 (19-60) | 28 (18-61) |
| Donor pregnancy |  |  |  |
| No | 893 (17) | 324 (16) | 569 (18) |
| Yes | 493 (10) | 187 (9) | 306 (10) |
| Male | 3708 (73) | 1514 (75) | 2194 (71) |
| Missing | 12 (<1) | 5 (<1) | 7 (<1) |
| Karnofsky Performance Score |  |  |  |
| Oct-80 | 2023 (40) | 769 (38) | 1254 (41) |
| 90-100 | 3015 (59) | 1230 (61) | 1785 (58) |
| Missing | 68 (1) | 31 (2) | 37 (1) |
| GVHD prophylaxis |  |  |  |
| Tacrolimus + MMF +- others | 739 (14) | 364 (18) | 375 (12) |
| Tacrolimus + MTX +- others (except MMF) | 3096 (61) | 1228 (60) | 1868 (61) |
| Tacrolimus + others (except MTX, MMF) | 400 (8) | 33 (2) | 367 (12) |
| Tacrolimus alone | 131 (3) | 119 (6) | 12 (<1) |
| CSA + MMF +- other(s) (except post-CY) | 332 (7) | 139 (7) | 193 (6) |
| CSA + MTX +- other(s) (except MMF, post-CY) | 301 (6) | 99 (5) | 202 (7) |
| CSA + other(s) (except MTX, MMF, post-CY) | 18 (<1) | 11 (1) | 7 (<1) |
| CSA alone | 19 (<1) | 7 (<1) | 12 (<1) |
| Other GVHD prophylaxis | 70 (1) | 30 (1) | 40 (1) |
| Graft type |  |  |  |
| Marrow | 885 (17) | 373 (18) | 512 (17) |
| PBSC | 4221 (83) | 1657 (82) | 2564 (83) |
| HCT-CI |  |  |  |
| 0 | 1166 (23) | 503 (25) | 663 (22) |
| 1 | 669 (13) | 286 (14) | 383 (12) |
| 2 | 788 (15) | 317 (16) | 471 (15) |
| 3+ | 2481 (49) | 923 (45) | 1558 (51) |
| NA/f2400 missing | 1 (<1) | 0 (<1) | 1 (<1) |
| Missing | 1 (<1) | 1 (<1) | 0 (<1) |
| Prior autologous transplant |  |  |  |
| No | 5022 (98) | 1997 (98) | 3025 (98) |
| Yes | 84 (2) | 33 (2) | 51 (2) |
| Conditioning regimen intensity |  |  |  |
| Myeloablative | 2710 (53) | 1032 (51) | 1678 (55) |
| RIC/NMA | 2380 (47) | 995 (49) | 1385 (45) |
| Missing | 16 (<1) | 3 (<1) | 13 (<1) |
| Donor/recipient sex match |  |  |  |
| Male/Male | 2272 (44) | 929 (46) | 1343 (44) |
| Male/Female | 1436 (28) | 585 (29) | 851 (28) |
| Female/Male | 636 (12) | 255 (13) | 381 (12) |
| Female/Female | 760 (15) | 260 (13) | 500 (16) |
| Missing | 2 (<1) | 1 (<1) | 1 (<1) |
| Donor/Recipient race match |  |  |  |
| Matched | 3 199 (63) | 1281 (63) | 1918 (62) |
| Mismatched | 331 (6) | 142 (7) | 189 (6) |
| Missing | 1576 (31) | 607 (30) | 969 (32) |
| Donor/recipient CMV serostatus |  |  |  |
| Positive/Positive | 1348 (26) | 572 (28) | 776 (25) |
| Positive/Negative | 470 (9) | 179 (9) | 291 (9) |
| Negative/Positive | 1843 (36) | 760 (37) | 1083 (35) |
| Negative/Negative | 1415 (28) | 509 (25) | 906 (29) |
| Missing | 30 (1) | 10 (<1) | 20 (1) |
| Year of transplant |  |  |  |
| 2008 | 70 (1) | 18 (1) | 52 (2) |
| 2009 | 196 (4) | 68 (3) | 128 (4) |
| 2010 | 425 (8) | 166 (8) | 259 (8) |
| 2011 | 480 (9) | 205 (10) | 275 (9) |
| 2012 | 570 (11) | 228 (11) | 342 (11) |
| 2013 | 548 (11) | 210 (10) | 338 (11) |
| 2014 | 690 (14) | 295 (15) | 395 (13) |
| 2015 | 1164 (23) | 453 (22) | 711 (23) |
| 2016 | 780 (15) | 325 (16) | 455 (15) |
| 2017 | 183 (4) | 62 (3) | 121 (4) |
| Indicator of death |  |  |  |
| Censoring | 2430 (48) | 960 (47) | 1470 (48) |
| Event | 2676 (52) | 1070 (53) | 1606 (52) |
| Missing | 0 (<1) | 0 (<1) | 0 (<1) |

**Supplemental Table 2.** Hazards of Grades II-IV Acute GVHD comparing 1 DPB1 mismatch (restricting to 10/10 matching on other alleles) versus complete matching (12 for 12). The results are split first by the deplete and replete cohorts, then a comparison across DPB1 mismatches categories of any, TCE permissiveness, expression, and evolutionary clade. For the evolutionary model DP5vsDP2 corresponds to a patient with DP5 versus a donor with DP2 and is flipped for DP2vsDP5.

| *Cohort* | *Category*<br>*DPB1* | *Mismatch*<br>*Type* | *Matches* | *Mismatches* | *Hazard Ratio*<br>*(Confidence Int.)* | *P*<br>*Value* |
| --- | --- | --- | --- | --- | --- | --- |
| *Deplete* | Any | Any | 228 | 922 | 1.18 (0.93, 1.51) | 0.17 |
|  | TCE | Non-Perm | 228 | 314 | 1.28 (0.95, 1.71) | 0.10 |
|  |  | Permissive | 228 | 608 | 1.14 (0.88, 1.48) | 0.32 |
|  | Expression | High | 228 | 397 | 1.40 (1.07, 1.84) | 0.016 |
|  |  | Low | 228 | 521 | 1.04 (0.79, 1.35) | 0.79 |
|  | Clade | DP5vsDP2 | 228 | 269 | 1.38 (1.02, 1.86) | 0.035 |
|  |  | DP2vsDP5 | 228 | 239 | 0.96 (0.69, 1.32) | 0.80 |
|  |  | Within | 228 | 414 | 1.21 (0.92, 1.59) | 0.18 |
| *Replete* | Any | Any | 361 | 1312 | 1.17 (0.98, 1.41) | 0.089 |
|  | TCE | Non-Perm | 361 | 455 | 1.23 (1.00, 1.53) | 0.054 |
|  |  | Permissive | 361 | 857 | 1.14 (0.94, 1.38) | 0.18 |
|  | Expression | High | 361 | 567 | 1.32 (1.07, 1.62) | 0.0082 |
|  |  | Low | 361 | 740 | 1.07 (0.88, 1.30) | 0.52 |
|  | Clade | DP5vsDP2 | 361 | 370 | 1.36 (1.09, 1.69) | 0.0068 |
|  |  | DP2vsDP5 | 361 | 299 | 1.04 (0.82, 1.33) | 0.73 |
|  |  | Within | 361 | 643 | 1.13 (0.92, 1.38) | 0.24 |

**Supplemental Table 3.** Hazards of overall survival comparing 1 DPB1 mismatch (restricting to 10/10 matching on other alleles) versus complete matching (12/12).

| *Cohort* | *Category*<br>*DPB1* | *Mismatch*<br>*Type* | *Matches* | *Mismatches* | *Hazard Ratio*<br>*(Confidence Int.)* | *P*<br>*Value* |
| --- | --- | --- | --- | --- | --- | --- |
| *Deplete* | Any | Any | 243 | 964 | 1.13 (0.92, 1.38) | 0.24 |
|  | TCE | Non-Perm | 243 | 327 | 1.19 (0.93, 1.53) | 0.17 |
|  |  | Permissive | 243 | 637 | 1.10 (0.89, 1.36) | 0.38 |
|  | Expression | High | 243 | 412 | 1.19 (0.94, 1.51) | 0.15 |
|  |  | Low | 243 | 548 | 1.08 (0.87, 1.35) | 0.48 |
|  | Clade | DP5vsDP2 | 243 | 280 | 1.18 (0.91, 1.54) | 0.21 |
|  |  | DP2vsDP5 | 243 | 253 | 1.11 (0.85, 1.44) | 0.44 |
|  |  | Within | 243 | 431 | 1.11 (0.88, 1.39) | 0.39 |
| *Replete* | Any | Any | 386 | 1406 | 0.94 (0.80, 1.10) | 0.44 |
|  | TCE | Non-Perm | 386 | 494 | 1.03 (0.85, 1.24) | 0.79 |
|  |  | Permissive | 386 | 912 | 0.90 (0.76, 1.06) | 0.21 |
|  | Expression | High | 386 | 604 | 0.97 (0.81, 1.16) | 0.73 |
|  |  | Low | 386 | 797 | 0.92 (0.77, 1.09) | 0.32 |
|  | Clade | DP5vsDP2 | 386 | 397 | 0.98 (0.81, 1.20) | 0.87 |
|  |  | DP2vsDP5 | 386 | 325 | 0.97 (0.79, 1.20) | 0.79 |
|  |  | Within | 386 | 684 | 0.90 (0.75, 1.08) | 0.26 |

**Supplemental Table 4.** Hazards of treatment related mortality comparing 1 DPB1 mismatch (restricting to 10/10 matching on other alleles) versus complete matching (12/12).

| *Cohort* | *Category*<br>*DPB1* | *Mismatch*<br>*Type* | *Matches* | *Mismatches* | *Hazard Ratio*<br>*(Confidence Int.)* | *P*<br>*Value* |
| --- | --- | --- | --- | --- | --- | --- |
| *Deplete* | Any | Any | 240 | 948 | 1.17 (0.85, 1.60) | 0.34 |
|  | TCE | Non-Perm | 240 | 322 | 1.16 (0.79, 1.70) | 0.45 |
|  |  | Permissive | 240 | 626 | 1.17 (0.84, 1.64) | 0.34 |
|  | Expression | High | 240 | 407 | 1.31 (0.92, 1.88) | 0.14 |
|  |  | Low | 240 | 537 | 1.06 (0.75, 1.49) | 0.74 |
|  | Clade | DP5vsDP2 | 240 | 277 | 1.26 (0.85, 1.87) | 0.24 |
|  |  | DP2vsDP5 | 240 | 247 | 1.03 (0.69, 1.54) | 0.89 |
|  |  | Within | 240 | 424 | 1.20 (0.84, 1.71) | 0.32 |
| *Replete* | Any | Any | 382 | 1392 | 0.86 (0.70, 1.07) | 0.17 |
|  | TCE | Non-Perm | 382 | 489 | 0.97 (0.75, 1.26) | 0.84 |
|  |  | Permissive | 382 | 903 | 0.80 (0.64, 1.01) | 0.060 |
|  | Expression | High | 382 | 596 | 0.91 (0.71, 1.16) | 0.43 |
|  |  | Low | 382 | 791 | 0.83 (0.66, 1.05) | 0.12 |
|  | Clade | DP5vsDP2 | 382 | 392 | 0.95 (0.72, 1.24) | 0.70 |
|  |  | DP2vsDP5 | 382 | 321 | 0.94 (0.70, 1.25) | 0.65 |
|  |  | Within | 382 | 679 | 0.78 (0.61, 0.99) | 0.044 |

**Supplemental Table 5.**
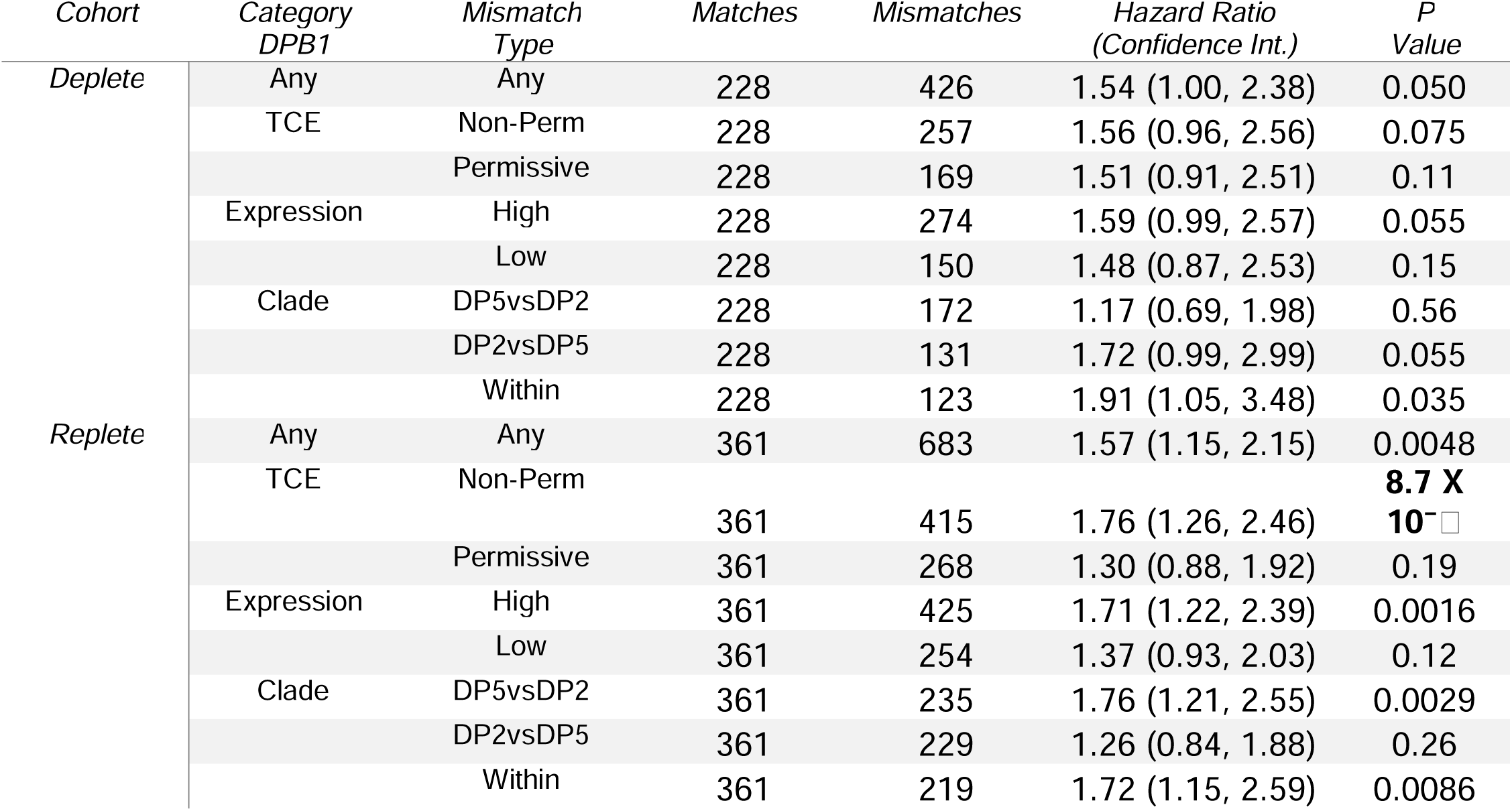
Hazards of Grades 3-4 Acute GVHD comparing 2 DPB1 mismatches (restricting to 10/10 matching on other alleles) versus complete matching (12/12).

**Supplemental Table 6.**
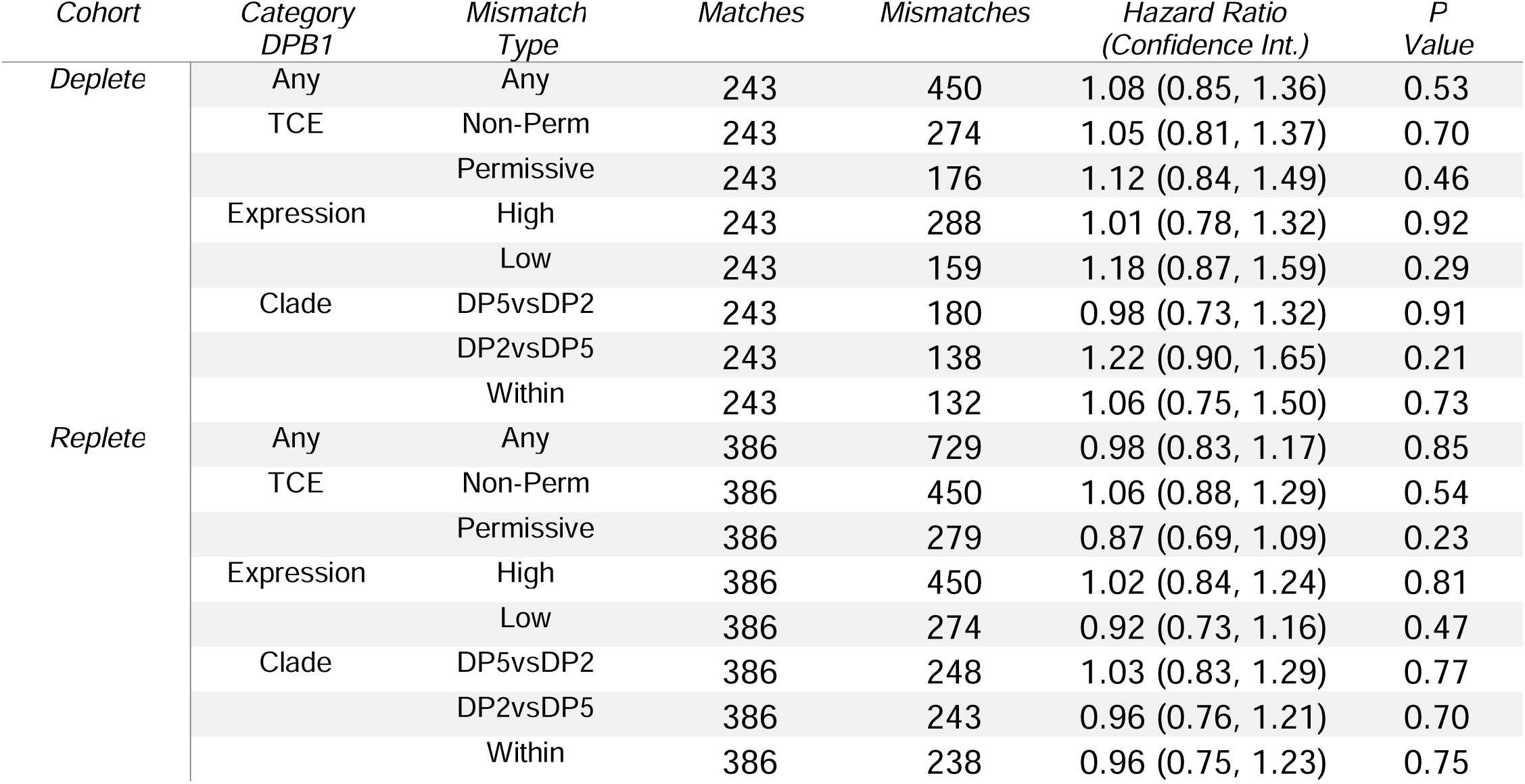
Hazards of overall survival Acute GVHD comparing 2 DPB1 mismatches (restricting to 10/10 matching on other alleles) versus complete matching (12/12).

| *Cohort* | *Category*<br>*DPB1* | *Mismatch*<br>*Type* | *Matches* | *Mismatches* | *Hazard Ratio*<br>*(Confidence Int.)* | *P*<br>*Value* |
| --- | --- | --- | --- | --- | --- | --- |
| *Deplete* | Any | Any | 243 | 450 | 1.08 (0.85, 1.36) | 0.53 |
|  | TCE | Non-Perm | 243 | 274 | 1.05 (0.81, 1.37) | 0.70 |
|  |  | Permissive | 243 | 176 | 1.12 (0.84, 1.49) | 0.46 |
|  | Expression | High | 243 | 288 | 1.01 (0.78, 1.32) | 0.92 |
|  |  | Low | 243 | 159 | 1.18 (0.87, 1.59) | 0.29 |
|  | Clade | DP5vsDP2 | 243 | 180 | 0.98 (0.73, 1.32) | 0.91 |
|  |  | DP2vsDP5 | 243 | 138 | 1.22 (0.90, 1.65) | 0.21 |
|  |  | Within | 243 | 132 | 1.06 (0.75, 1.50) | 0.73 |
| *Replete* | Any | Any | 386 | 729 | 0.98 (0.83, 1.17) | 0.85 |
|  | TCE | Non-Perm | 386 | 450 | 1.06 (0.88, 1.29) | 0.54 |
|  |  | Permissive | 386 | 279 | 0.87 (0.69, 1.09) | 0.23 |
|  | Expression | High | 386 | 450 | 1.02 (0.84, 1.24) | 0.81 |
|  |  | Low | 386 | 274 | 0.92 (0.73, 1.16) | 0.47 |
|  | Clade | DP5vsDP2 | 386 | 248 | 1.03 (0.83, 1.29) | 0.77 |
|  |  | DP2vsDP5 | 386 | 243 | 0.96 (0.76, 1.21) | 0.70 |
|  |  | Within | 386 | 238 | 0.96 (0.75, 1.23) | 0.75 |

**Supplemental Table 7.** Hazards of treatment related mortality comparing 2 DPB1 mismatches (restricting to 10/10 matching on other alleles) versus complete matching (12/12).

| *Cohort* | *Category*<br>*DPB1* | *Mismatch*<br>*Type* | *Matches* | *Mismatches* | *Hazard Ratio*<br>*(Confidence Int.)* | *P*<br>*Value* |
| --- | --- | --- | --- | --- | --- | --- |
| *Deplete* | Any | Any | 240 | 446 | 1.45 (1.03, 2.03) | 0.032 |
|  | TCE | Non-Perm | 240 | 271 | 1.42 (0.98, 2.08) | 0.067 |
|  |  | Permissive | 240 | 175 | 1.49 (0.99, 2.25) | 0.054 |
|  | Expression | High | 240 | 285 | 1.34 (0.92, 1.95) | 0.13 |
|  |  | Low | 240 | 158 | 1.68 (1.12, 2.53) | 0.012 |
|  | Clade | DP5vsDP2 | 240 | 178 | 1.20 (0.79, 1.84) | 0.40 |
|  |  | DP2vsDP5 | 240 | 136 | 1.63 (1.07, 2.49) | 0.024 |
|  |  | Within | 240 | 132 | 1.60 (1.01, 2.53) | 0.047 |
| *Replete* | Any | Any | 382 | 715 | 1.00 (0.79, 1.26) | 0.99 |
|  | TCE | Non-Perm | 382 | 442 | 1.12 (0.87, 1.45) | 0.38 |
|  |  | Permissive | 382 | 273 | 0.82 (0.59, 1.12) | 0.21 |
|  | Expression | High | 382 | 445 | 1.07 (0.83, 1.38) | 0.61 |
|  |  | Low | 382 | 265 | 0.89 (0.65, 1.21) | 0.46 |
|  | Clade | DP5vsDP2 | 382 | 244 | 0.97 (0.72, 1.32) | 0.86 |
|  |  | DP2vsDP5 | 382 | 235 | 0.86 (0.63, 1.18) | 0.36 |
|  |  | Within | 382 | 236 | 1.17 (0.85, 1.59) | 0.33 |

**Supplemental Table 8.**
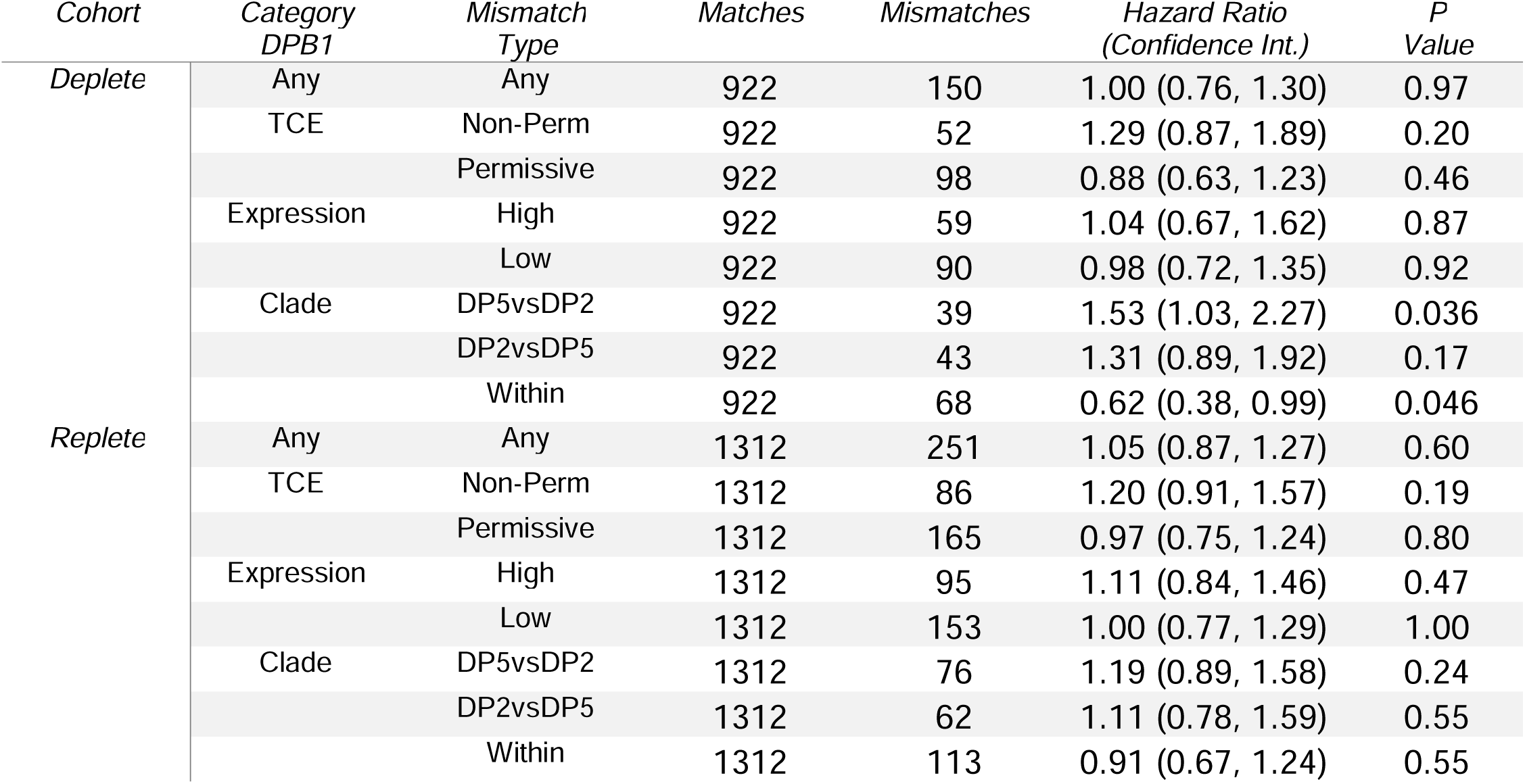
Hazards of acute GVHD 2-4 between additive model of noncoding mismatches with one DPB1 mismatches versus baseline of 10/10 matching with one DPB1 mismatch.

| *Cohort* | *Category*<br>*DPB1* | *Mismatch*<br>*Type* | *Matches* | *Mismatches* | *Hazard Ratio*<br>*(Confidence Int.)* | *P*<br>*Value* |
| --- | --- | --- | --- | --- | --- | --- |
| *Deplete* | Any | Any | 922 | 150 | 1.00 (0.76, 1.30) | 0.97 |
|  | TCE | Non-Perm | 922 | 52 | 1.29 (0.87, 1.89) | 0.20 |
|  |  | Permissive | 922 | 98 | 0.88 (0.63, 1.23) | 0.46 |
|  | Expression | High | 922 | 59 | 1.04 (0.67, 1.62) | 0.87 |
|  |  | Low | 922 | 90 | 0.98 (0.72, 1.35) | 0.92 |
|  | Clade | DP5vsDP2 | 922 | 39 | 1.53 (1.03, 2.27) | 0.036 |
|  |  | DP2vsDP5 | 922 | 43 | 1.31 (0.89, 1.92) | 0.17 |
|  |  | Within | 922 | 68 | 0.62 (0.38, 0.99) | 0.046 |
| *Replete* | Any | Any | 1312 | 251 | 1.05 (0.87, 1.27) | 0.60 |
|  | TCE | Non-Perm | 1312 | 86 | 1.20 (0.91, 1.57) | 0.19 |
|  |  | Permissive | 1312 | 165 | 0.97 (0.75, 1.24) | 0.80 |
|  | Expression | High | 1312 | 95 | 1.11 (0.84, 1.46) | 0.47 |
|  |  | Low | 1312 | 153 | 1.00 (0.77, 1.29) | 1.00 |
|  | Clade | DP5vsDP2 | 1312 | 76 | 1.19 (0.89, 1.58) | 0.24 |
|  |  | DP2vsDP5 | 1312 | 62 | 1.11 (0.78, 1.59) | 0.55 |
|  |  | Within | 1312 | 113 | 0.91 (0.67, 1.24) | 0.55 |

**Supplemental Table 9.**
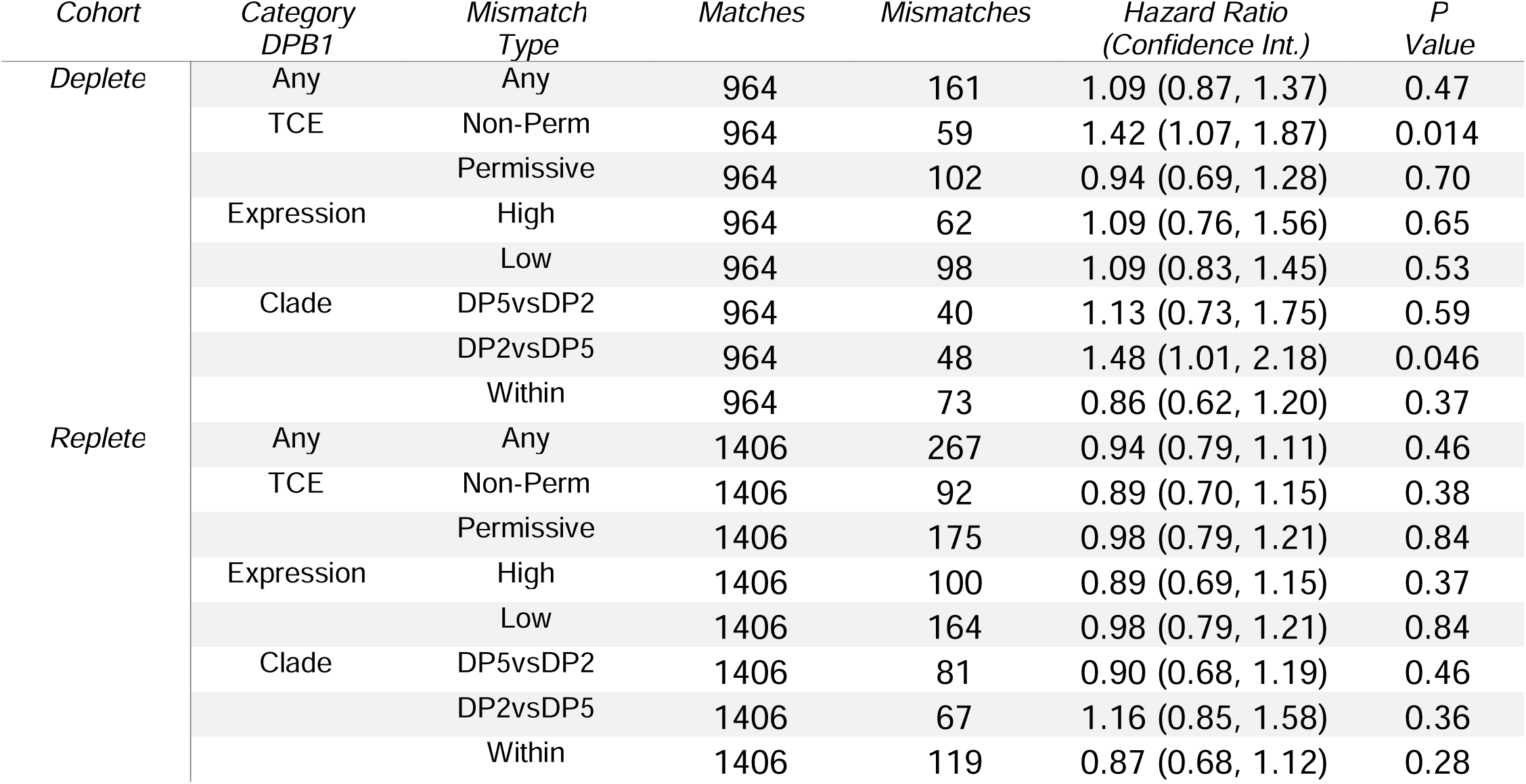
Hazards of overall survival between additive model of noncoding mismatches with one DPB1 mismatches versus baseline of 10/10 matching with one DPB1 mismatch.

| *Cohort* | *Category*<br>*DPB1* | *Mismatch*<br>*Type* | *Matches* | *Mismatches* | *Hazard Ratio*<br>*(Confidence Int.)* | *P*<br>*Value* |
| --- | --- | --- | --- | --- | --- | --- |
| *Deplete* | Any | Any | 964 | 161 | 1.09 (0.87, 1.37) | 0.47 |
|  | TCE | Non-Perm | 964 | 59 | 1.42 (1.07, 1.87) | 0.014 |
|  |  | Permissive | 964 | 102 | 0.94 (0.69, 1.28) | 0.70 |
|  | Expression | High | 964 | 62 | 1.09 (0.76, 1.56) | 0.65 |
|  |  | Low | 964 | 98 | 1.09 (0.83, 1.45) | 0.53 |
|  | Clade | DP5vsDP2 | 964 | 40 | 1.13 (0.73, 1.75) | 0.59 |
|  |  | DP2vsDP5 | 964 | 48 | 1.48 (1.01, 2.18) | 0.046 |
|  |  | Within | 964 | 73 | 0.86 (0.62, 1.20) | 0.37 |
| *Replete* | Any | Any | 1406 | 267 | 0.94 (0.79, 1.11) | 0.46 |
|  | TCE | Non-Perm | 1406 | 92 | 0.89 (0.70, 1.15) | 0.38 |
|  |  | Permissive | 1406 | 175 | 0.98 (0.79, 1.21) | 0.84 |
|  | Expression | High | 1406 | 100 | 0.89 (0.69, 1.15) | 0.37 |
|  |  | Low | 1406 | 164 | 0.98 (0.79, 1.21) | 0.84 |
|  | Clade | DP5vsDP2 | 1406 | 81 | 0.90 (0.68, 1.19) | 0.46 |
|  |  | DP2vsDP5 | 1406 | 67 | 1.16 (0.85, 1.58) | 0.36 |
|  |  | Within | 1406 | 119 | 0.87 (0.68, 1.12) | 0.28 |

**Supplemental Table 10.** Hazards of treatment related mortality between additive model of noncoding mismatches with one DPB1 mismatches versus baseline of 10/10 matching with one DPB1 mismatch.

| *Cohort* | *Category*<br>*DPB1* | *Mismatch*<br>*Type* | *Matches* | *Mismatches* | *Hazard Ratio*<br>*(Confidence Int.)* | *P*<br>*Value* |
| --- | --- | --- | --- | --- | --- | --- |
| *Deplete* | Any | Any | 948 | 156 | 1.27 (0.92, 1.77) | 0.15 |
|  | TCE | Non-Perm | 948 | 59 | 1.57 (0.95, 2.59) | 0.079 |
|  |  | Permissive | 948 | 97 | 1.15 (0.77, 1.72) | 0.49 |
|  | Expression | High | 948 | 61 | 1.02 (0.55, 1.87) | 0.96 |
|  |  | Low | 948 | 94 | 1.39 (0.96, 2.01) | 0.082 |
|  | Clade | DP5vsDP2 | 948 | 40 | 1.34 (0.70, 2.54) | 0.38 |
|  |  | DP2vsDP5 | 948 | 47 | 2.20 (1.32, 3.65) | 0.0023 |
|  |  | Within | 948 | 69 | 0.74 (0.44, 1.25) | 0.26 |
| *Replete* | Any | Any | 1392 | 264 | 0.91 (0.72, 1.15) | 0.42 |
|  | TCE | Non-Perm | 1392 | 92 | 0.86 (0.61, 1.22) | 0.40 |
|  |  | Permissive | 1392 | 172 | 0.96 (0.71, 1.29) | 0.76 |
|  | Expression | High | 1392 | 100 | 0.92 (0.67, 1.26) | 0.59 |
|  |  | Low | 1392 | 161 | 0.87 (0.64, 1.19) | 0.39 |
|  | Clade | DP5vsDP2 | 1392 | 81 | 0.91 (0.64, 1.28) | 0.57 |
|  |  | DP2vsDP5 | 1392 | 66 | 0.94 (0.58, 1.52) | 0.80 |
|  |  | Within | 1392 | 117 | 0.91 (0.64, 1.31) | 0.62 |

**Supplemental Table 11.** Comparative Analysis: Exclusion of Noncoding Sequences and Its Influence on Hazard Ratio Estimates for Treatment-Related Mortality in DPB1 Mismatch Categories. The hazard ratio differences (HR Diff), and p-value differences (p-value) compare the relative values using only coding versus the models fit with coding and noncoding data. This corresponds to the same data plotted in Figure 3 and Supplemental Figure 2.

| Cohort | DPB1 Set | Mismatch Type | Only Coding (2 Field Typing) |  |  |  | Coding and noncoding (4 Field Typing) |  |  |  | HR Diff | P-value Diff |
| --- | --- | --- | --- | --- | --- | --- | --- | --- | --- | --- | --- | --- |
|  |  |  | Matches | Mis-matches | Hazard Ratio (Conf. Int.) | P Value | Matches | Mis-matches | Hazard Ratio (Conf. Int.) | P Value |  |  |
| Deplete | Any | Any | 288 | 561 | 1.59 (1.17, 2.16) | 0.0030 | 240 | 446 | 1.45 (1.03, 2.03) | 0.032 | 0.14 | -0.029 |
|  | TCE | Non-Perm | 288 | 347 | 1.55 (1.11, 2.17) | 0.010 | 240 | 271 | 1.42 (0.98, 2.08) | 0.067 | 0.13 | -0.057 |
|  |  | Permissive | 288 | 214 | 1.66 (1.15, 2.38) | 0.0064 | 240 | 175 | 1.49 (0.99, 2.25) | 0.054 | 0.16 | -0.048 |
|  | Expression | High | 288 | 355 | 1.41 (1.02, 1.97) | 0.040 | 240 | 285 | 1.34 (0.92, 1.95) | 0.13 | 0.077 | -0.093 |
|  |  | Low | 288 | 202 | 1.93 (1.33, 2.79) | 5.0 X 10<sup>-4</sup> | 240 | 158 | 1.68 (1.12, 2.53) | 0.012 | 0.25 | -0.012 |
|  | Clade | DP5vsDP2 | 288 | 219 | 1.28 (0.88, 1.86) | 0.20 | 240 | 178 | 1.20 (0.79, 1.84) | 0.40 | 0.073 | -0.19 |
|  |  | DP2vsDP5 | 288 | 180 | 2.10 (1.44, 3.05) | 1.0 X 10<sup>-4</sup> | 240 | 136 | 1.63 (1.07, 2.49) | 0.024 | 0.47 | -0.024 |
|  |  | Within | 288 | 162 | 1.49 (1.00, 2.21) | 0.050 | 240 | 132 | 1.60 (1.01, 2.53) | 0.047 | -0.11 | 0.0022 |
| Replete | Any | Any | 468 | 864 | 0.92 (0.74, 1.16) | 0.49 | 382 | 715 | 1.00 (0.79, 1.26) | 0.99 | -0.075 | -0.50 |
|  | TCE | Non-Perm | 468 | 534 | 1.04 (0.81, 1.34) | 0.73 | 382 | 442 | 1.12 (0.87, 1.45) | 0.38 | -0.077 | 0.35 |
|  |  | Permissive | 468 | 329 | 0.75 (0.55, 1.02) | 0.071 | 382 | 273 | 0.82 (0.59, 1.12) | 0.21 | -0.064 | -0.14 |
|  | Expression | High | 468 | 535 | 1.01 (0.79, 1.29) | 0.93 | 382 | 445 | 1.07 (0.83, 1.38) | 0.61 | -0.057 | 0.31 |
|  |  | Low | 468 | 323 | 0.80 (0.58, 1.09) | 0.15 | 382 | 265 | 0.89 (0.65, 1.21) | 0.46 | -0.093 | -0.30 |
|  | Clade | DP5vsDP2 | 468 | 289 | 0.97 (0.72, 1.30) | 0.82 | 382 | 244 | 0.97 (0.72, 1.32) | 0.86 | -0.0068 | -0.038 |
|  |  | DP2vsDP5 | 468 | 295 | 0.82 (0.60, 1.13) | 0.22 | 382 | 235 | 0.86 (0.63, 1.18) | 0.36 | -0.043 | -0.14 |
|  |  | Within | 468 | 280 | 0.99 (0.73, 1.35) | 0.96 | 382 | 236 | 1.17 (0.85, 1.59) | 0.33 | -0.17 | 0.63 |

**Supplemental Table 12.**
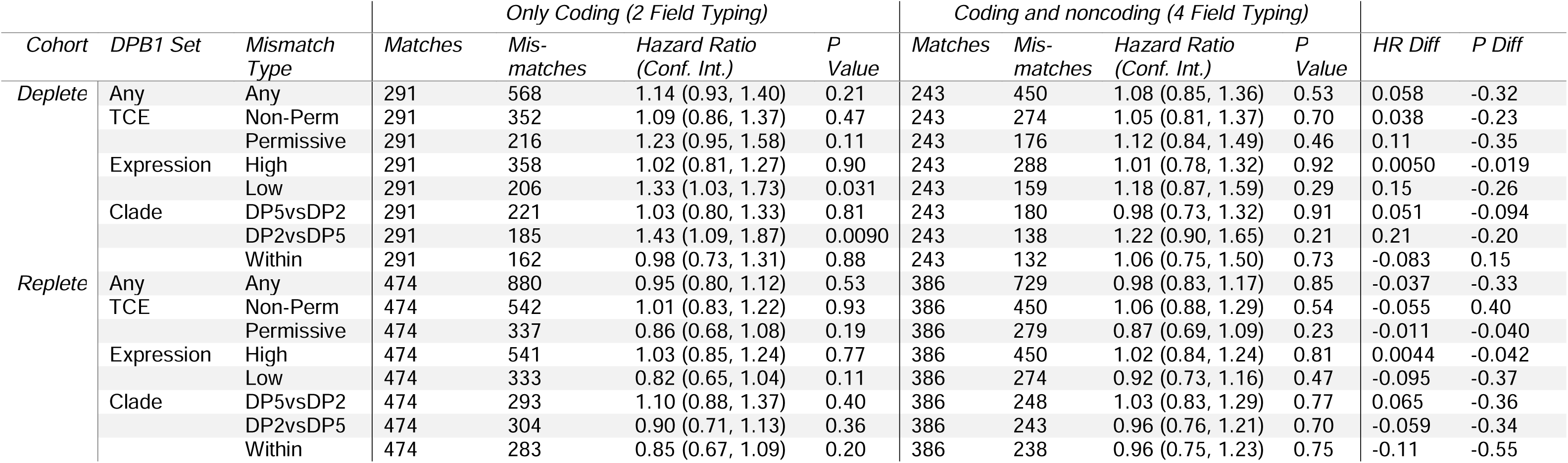
Comparative Analysis: Exclusion of Noncoding Sequences and Its Influence on Hazard Ratio Estimates for Overall Survival in DPB1 Mismatch Categories. The hazard ratio differences (HR Diff), and p-value differences (p-value) compare the relative values using only coding versus the models fit with coding and noncoding data. This corresponds to the same data plotted in Figure 3 and Supplemental Figure 2.

**Supplemental Figure 1.**
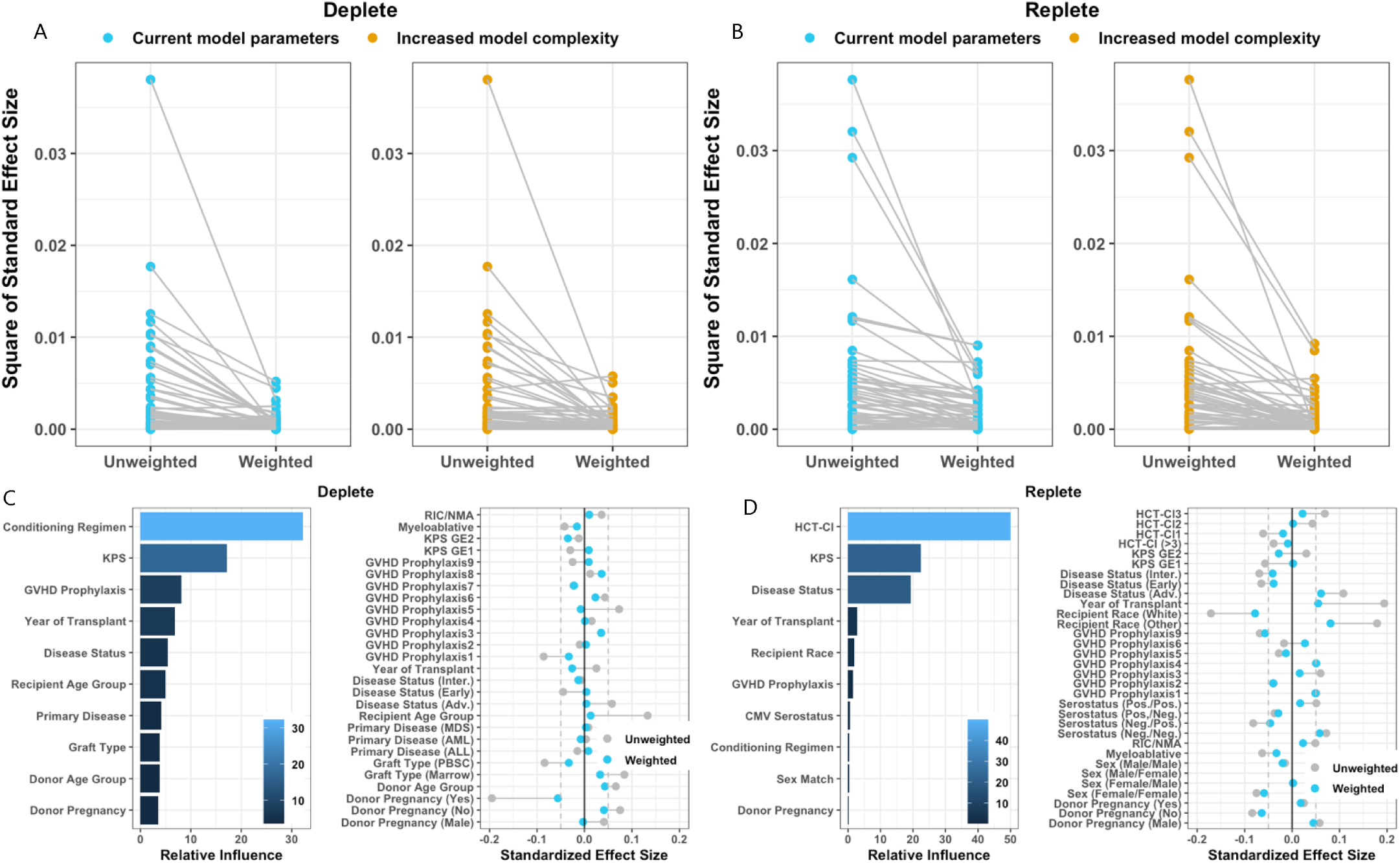
Controlling for possible confounders using gradient boosted model to estimate propensity of finding a perfect HLA match. To test how different HLA mismatches affect outcomes, we first consider the probability or propensity of achieving a perfect match to control for confounding. In this context, the propensity scores represent the likelihood of obtaining a UHR match (the primary exposure) based on the patient’s characteristics. The variables used to estimate these propensity scores include a range of covariates related to both patient and donor characteristics (see Supplemental Table 1). Before fitting the final model, we conducted a sensitivity analysis by fitting the GBM model with various input parameters for both the A) deplete and B) replete cohorts. In blue, for both A) and B), the GBM was applied to a simpler model with 5,000 trees, a shrinkage parameter of 0.01, an interaction depth of 3, and stopping criteria based on the absolute standardized mean difference. This was compared to a more complex model with 10,000 trees, a shrinkage parameter of 0.00325, and an increased depth of 5. Each dot represents a covariate and its relative squared standard effect on finding a perfect match, first unweighted and then weighted by propensity scores. Lines shifting downward indicate better balance in covariates. The simpler model (blue) produced results very similar to the more complex model (orange), so we selected the simpler model to avoid potential overfitting. After selecting the input parameters based on sensitivity analysis, we evaluated the effects of these features both before and after weighting to address imbalances in finding a perfect match in the C) deplete cohort and D) replete cohort. We observed that after weighting based on the model the effects of the features shifted toward zero (as shown by the blue dots), indicating improved overall balance of covariates.

**Supplemental Figure 2.**
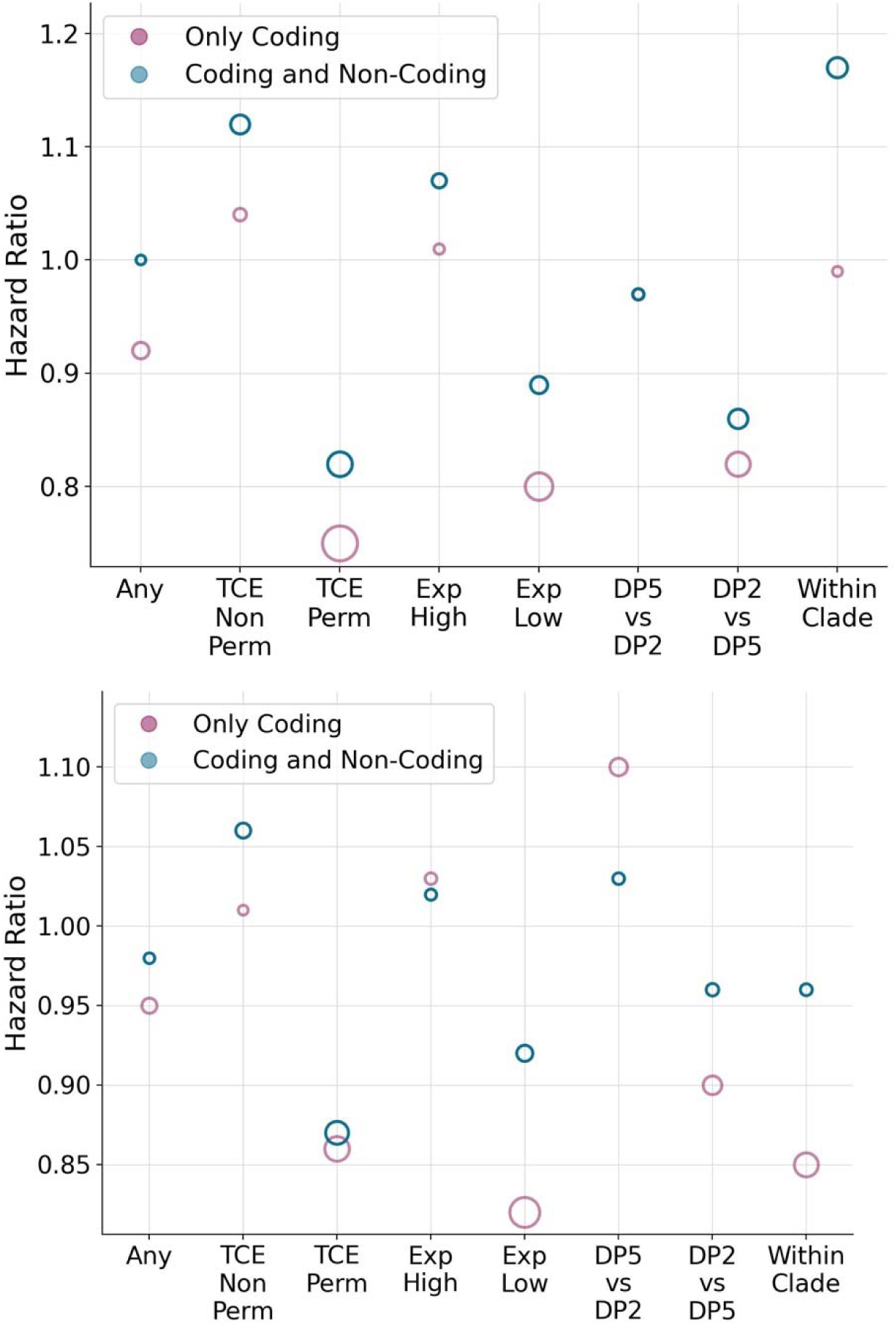
Hazards of treatment related mortality and overall survival in the context of two DPB1 mismatches and noncoding mismatches in Class I HLA for replete cohort. The plots show the hazard rates for A) TRM and B) overall survival comparing 12/12 matches verses 2 DPB1 MM (either TCE, expression, or clade) restricting to 10/10 matches when looking at only coding sequences (mimicking two field typing) versus coding and noncoding (4-field typing of HLA class I). The hazard rate dot sizes are proportional to the -log_10_(p-values) so, larger dots are more significant p-values and filled in dots are at a threshold of 0.01 or lower. Higher hazards indicate worse outcomes relative to baseline.

